# Mixed cytomegalovirus genotypes in HIV positive mothers show compartmentalization and distinct patterns of transmission to infants

**DOI:** 10.1101/2020.09.17.20196790

**Authors:** Juanita Pang, Jennifer A. Slyker, Sunando Roy, Josephine Bryant, Claire Atkinson, Juliana Cudini, Carey Farquhar, Paul Griffiths, James Kiarie, Sofia Morfopoulou, Alison C. Roxby, Helena Tutil, Rachel Williams, Soren Gantt, Richard A. Goldstein, Judith Breuer

## Abstract

Cytomegalovirus (CMV) is the most congenital infection (cCMVi), and is particularly common among infants born to HIV-infected women. Studies of cCMVi pathogenesis are complicated by the presence of multiple infecting maternal CMV strains, especially in HIV-positive women, and the large, recombinant CMV genome. Using newly-developed tools to reconstruct CMV haplotypes, we demonstrate anatomic CMV compartmentalization in five HIV-infected mothers, and identify the possibility of congenitally-transmitted genotypes in three of their infants. A single CMV strain was transmitted in each congenitally-infected case, and all were closely related to those that predominate in the cognate maternal cervix. Compared to non-transmitted strains, these congenitally-transmitted CMV strains showed statistically significant similarities in 19 genes associated with tissue-tropism and immunomodulation. In all infants, incident superinfections with distinct strains from breast milk were captured during follow-up. The results represent potentially important new insights into the virologic determinants of early CMV infection.

## Introduction

Human cytomegalovirus (CMV) is the commonest infectious cause of congenitally-acquired disability [1]. Between 0.2% and 2% of all live births have congenital CMV infection (cCMVi), and of these an estimated 15%-20% develop permanent sequelae ranging from sensorineural hearing loss to severe neurocognitive impairment [2, 3]. Maternal coinfection with HIV, even when mitigated by antiretroviral treatment, is associated with higher CMV viral loads in plasma, saliva, cervix and breast milk, and a greater risk of both congenital and postnatal CMV transmission [4-7]. Numerous studies have highlighted the negative health impacts of CMV on both HIV-infected and HIV-exposed uninfected (HEU) infants and children [8-10].

Primary maternal CMV infection during pregnancy confers a 30%-40% risk of transmission to the fetus [11]. Pre-existing maternal CMV immunity appears to reduce the risk of cCMVi, though it is clearly imperfect [12]. Due to their abundance in the community, over two-thirds of infants with cCMVi are born to seropositive women, and the overall risk of cCMVi is directly proportional to the maternal seroprevalence in a population [13]. Increasing evidence points to the importance of maternal CMV reinfection with new antigenic strains during pregnancy as a major risk factor for non-primary cCMVi [12, 14]. Evidence that household children may be a source of maternal reinfection provides additional support for this hypothesis [15, 16].

The CMV genome is the largest of the human herpesviruses. Regions of extensive sequence variability together with high levels of recombination between different strains results in high diversity for a DNA virus [17-19]. Individuals are often infected with multiple CMV strains. We have recently demonstrated that separate CMV haplotypes can be resolved from high-throughput sequencing (HTS) data [20]. This advance, by enabling tracking of individual genomes within mixed CMV infections, has already revealed the impact of mutation, recombination and selection in shaping the course of infection [20]. Here we apply these methods to CMV genomes sequenced from samples from five HIV-infected women and their infants that were collected between 1993 and 1998 originally for studies of maternal-infant HIV transmission [7]. By reconstructing genome-wide haplotypes from these longitudinal samples, we are able to examine the diversity of CMV shed by HIV-infected women and the specific genotypes that are transmitted in congenital and postnatal infections, and to reconstruct the likely chronology with which specific CMV variants were transmitted from mothers to infants.

## Results

### Participant characteristics, sampling, depth of sequencing

Details of the study cohort, follow-up, sample collection, and HIV and CMV infection status and transmission have been previously described [21-23]. Sufficient residual sample was available from the five families analysed here. To maximise the chance of recovering near full genomes, we selected samples reported in the original publication [22] to have > 10^3^ copies/ml, as this is the limit at which we generally can generate whole genomes from blood. Of the five mother-infant pairs analysed, four infants were HIV-exposed uninfected (HEU) (Infants 22, 123, 41, 14), and one was HIV-infected (Infant 12).

### CMV viral loads and sequencing

Cervical, breast milk, and blood viral loads, and time of sample collection for the five mother-infant pairs studied are shown in Fig. 1. The percentage of genome coverage and mean read depths are shown in Table 1. While breast milk samples had greater than 70% coverage at depths of 10x or more, the cervical and infant samples were of generally of lower depth, likely due to degradation of DNA due to the age and handling of the samples; genome coverage and mean de-duplicated read depth were directly related to actual CMV genome copy number present in the input material (Fig. S1). For all subsequent analysis, we removed samples with genome coverage of less than 20%. Fourteen of the remaining 20 cervical and baby samples had genome coverage above 70% and read depths of greater than 10x (Table 1).

**Table 1.**
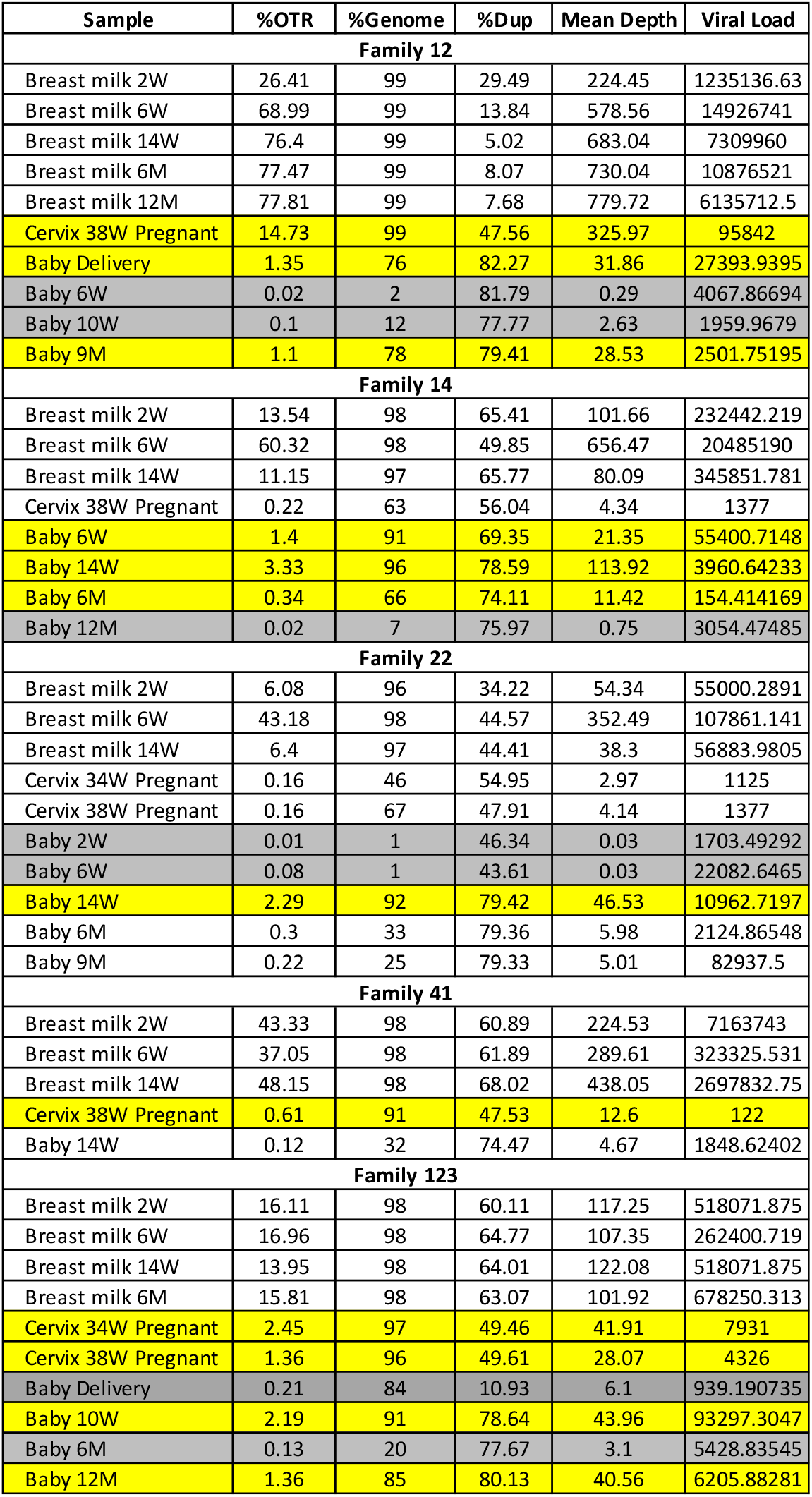
Sequencing characteristics for samples from each family. OTR: on target read; % Genome: % of genome coverage; % Dup: % of duplicated reads. Samples with genome coverages too low to be included in any analysis are shaded in grey. Cervical or baby samples with good coverage and read depth are highlighted in yellow.

**Fig. 1.**
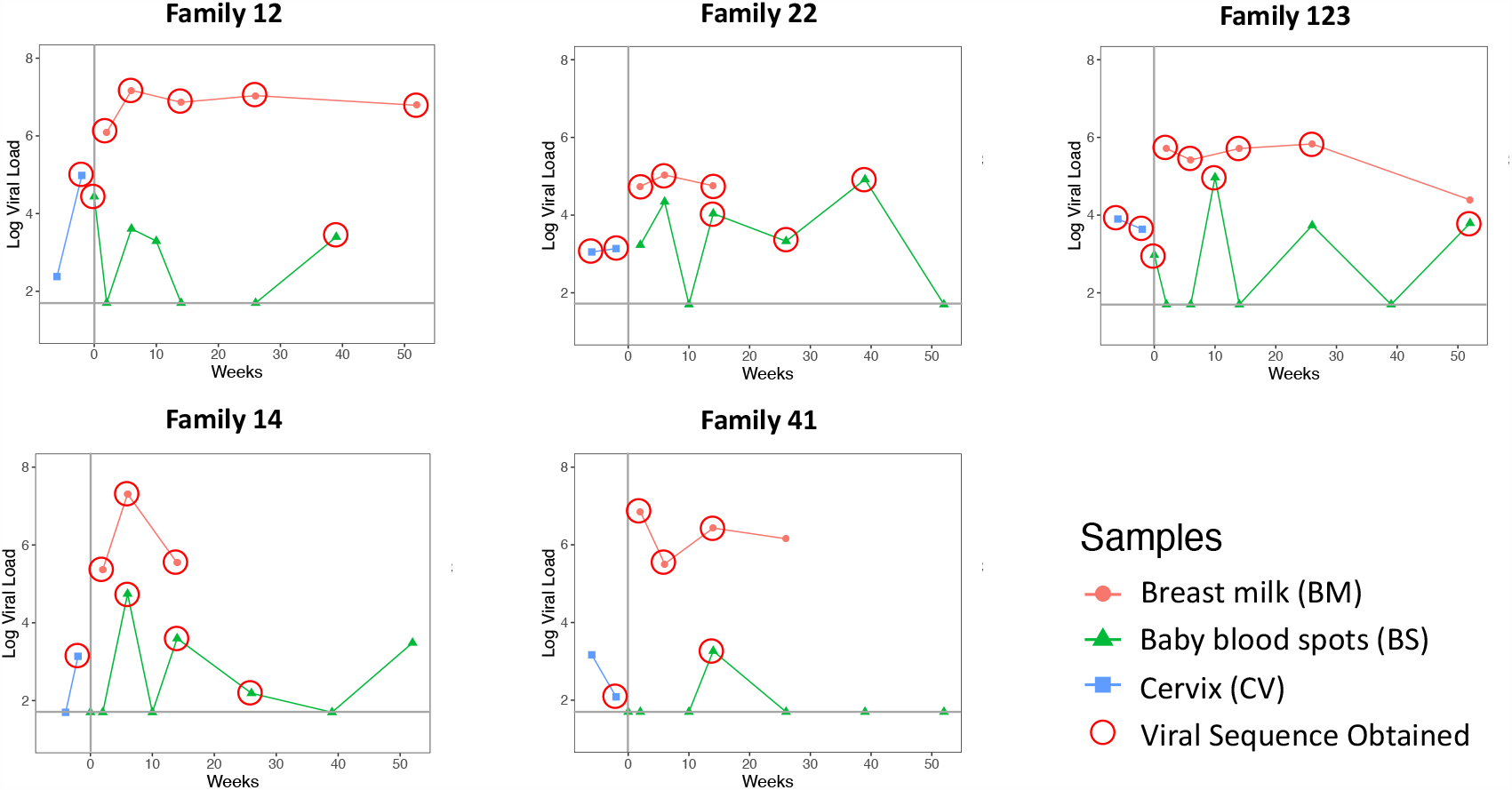
Viral loads of longitudinal samples for each family from breast milk (red), baby blood spots (green) and cervix (blue). Vertical grey line indicates date of delivery. Red circles indicate the samples that were submitted for whole genome sequencing.

### CMV genome sequence relatedness and diversity

We used multidimensional scaling to cluster CMV genomic sequences by nucleotide similarity (Fig. 2), as use of phylogenetic trees is problematic due to the high levels of CMV recombination. Sequences from families 12, 14 and 41 all clustered by family. Family 22 and 123 clustered in two distinct spaces, suggesting infection with more than one strain. In all five cases, the first sample from each infant (indicated by an arrow) clustered most closely with that of its mother, indicating the likelihood of recent maternal-infant transmission.

**Fig. 2.**
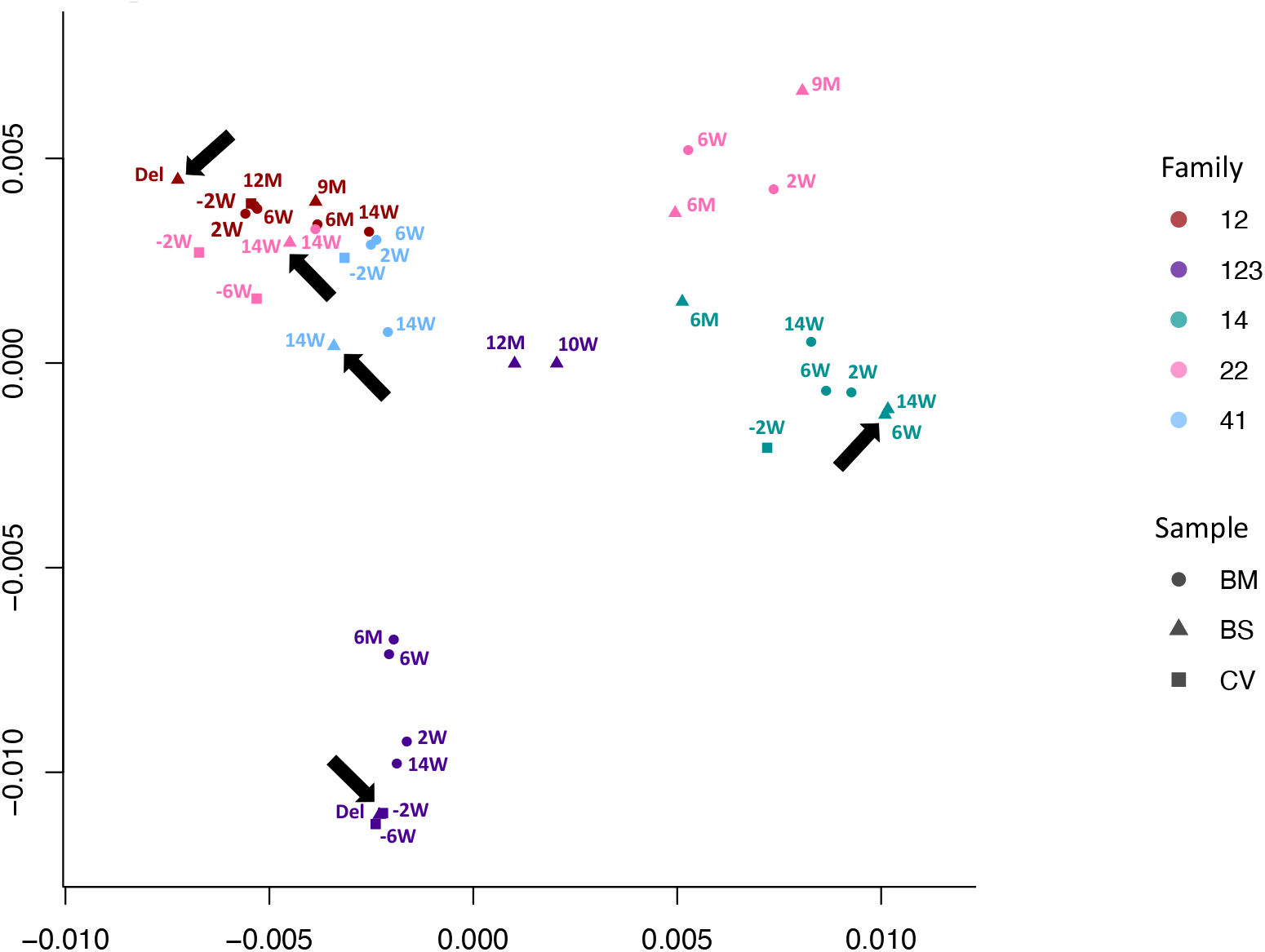
Multidimensional scaling showing clustering of consensus genome sequences for each sample by family. Arrows indicate that the first baby blood spot clusters with their own maternal sequences in all cases.

To further investigate the possibility of mixed infections, we calculated the within-sample nucleotide diversity, a metric that we have shown previously can be used as a proxy for the likelihood of mixed strain infections [20]. Fig. S2 shows that almost all the breast milk samples were highly diverse and therefore likely to contain multiple virus strains, a finding consistent with previous analyses of breast milk from HIV-infected women [24]. In contrast, the cervical and infant samples with the exception of one cervical sample from family 12, showed lower diversity. We used subsampling to demonstrate that computed nucleotide diversities are robust down to sequencing depths of >10 (Fig. S3). Low diversity was also observed in cervical and blood spots with higher coverage and read depths (Table 1).

### Reconstruction of individual haplotypes reveals CMV compartmentalization

To resolve the individual viral sequences (haplotypes) within each sample, we used our previously described method HaROLD [25]. Fig. 3 shows that haplotypes for each sample tended to cluster by family group albeit with clear evidence of distinct clusters even within a family e.g. family 22.

**Fig. 3.**
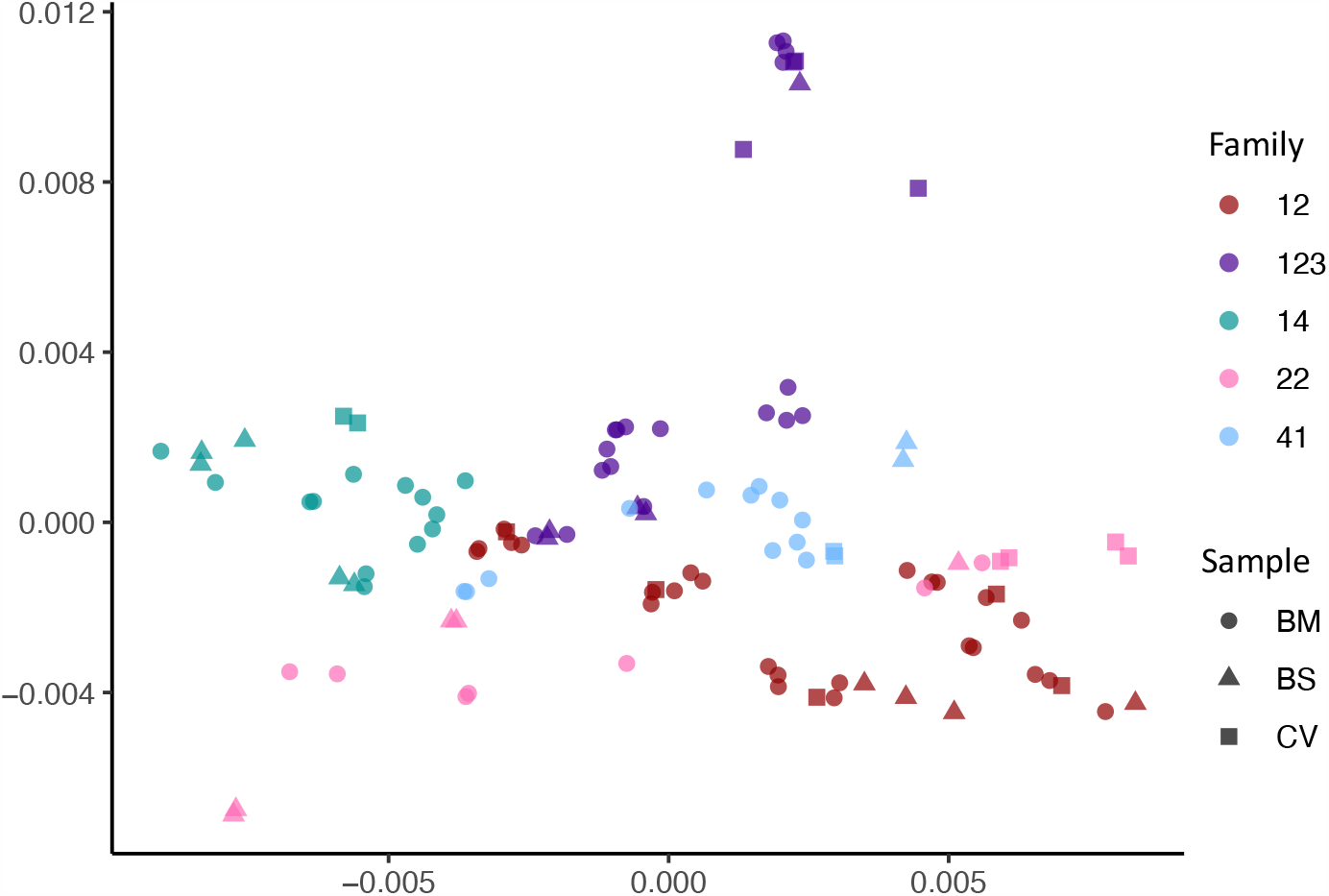
Multidimensional scaling showing clustering of haplotype sequences by family. Colours indicate the families, shapes indicate the types of sample.

The presence of mixed infections within a single family was supported by data showing that a subset of the sequence haplotypes within each family had pairwise distances as great as those between unrelated GenBank sequences (Fig. S4). Within-family phylogenetic analysis (Fig. S5) shows distinct clusters of the phylogenetically related sequence haplotypes recovered from breast milk, cervix and baby, likely to represent variants forming distinct viral strains (Fig. S5). Based on the distribution of pairwise distances (see Methods), we clustered similar haplotypes together into strains henceforth termed genotypes. In no cases did haplotypes from different families fulfil our clustering criterion confirming that haplotypes were not shared between unrelated families.

For ease of reference, genotypes were colored differently, with the genotype predominating in the first cervical sample of each family colored red (Fig. S5). Other genotypes were colored by their phylogenetic and pairwise distances from this genotype (Fig. S5). From our data, we identified at total of 26 genotypes with between 3 and 9 genotypes for each family (Fig. S5).

To elucidate the relationship between maternal and infant genotypes, we plotted the abundance of each within a sample over time (Fig. 4). All five mothers were infected with multiple genotypes in breast milk. In many cases genotypes within a single maternal sample were as genetically distant as unrelated database sequences, suggesting the presence of multiple distinct CMV strains (Fig. S5, Fig. 4). Relative genotype abundances present in breast milk changed over time. One unique genotype appeared in the breast milk of mother 22 at 6 weeks, disappearing from a subsequent sample (Fig. 4). This genotype was genetically distinct not only from other genotypes in family 22 but from genotypes in all other families, reducing the likelihood that it was a contaminant and may therefore have represented a new reinfection or reactivation of pre-existing latent infection. All cervical samples showed a single dominant genotype (Fig. 4), including mother 12, whose sample was more diverse and found to contain low levels of other genotypes. Overall, the data point to compartmentalization of CMV populations between cervix and breast milk.

**Fig. 4.**
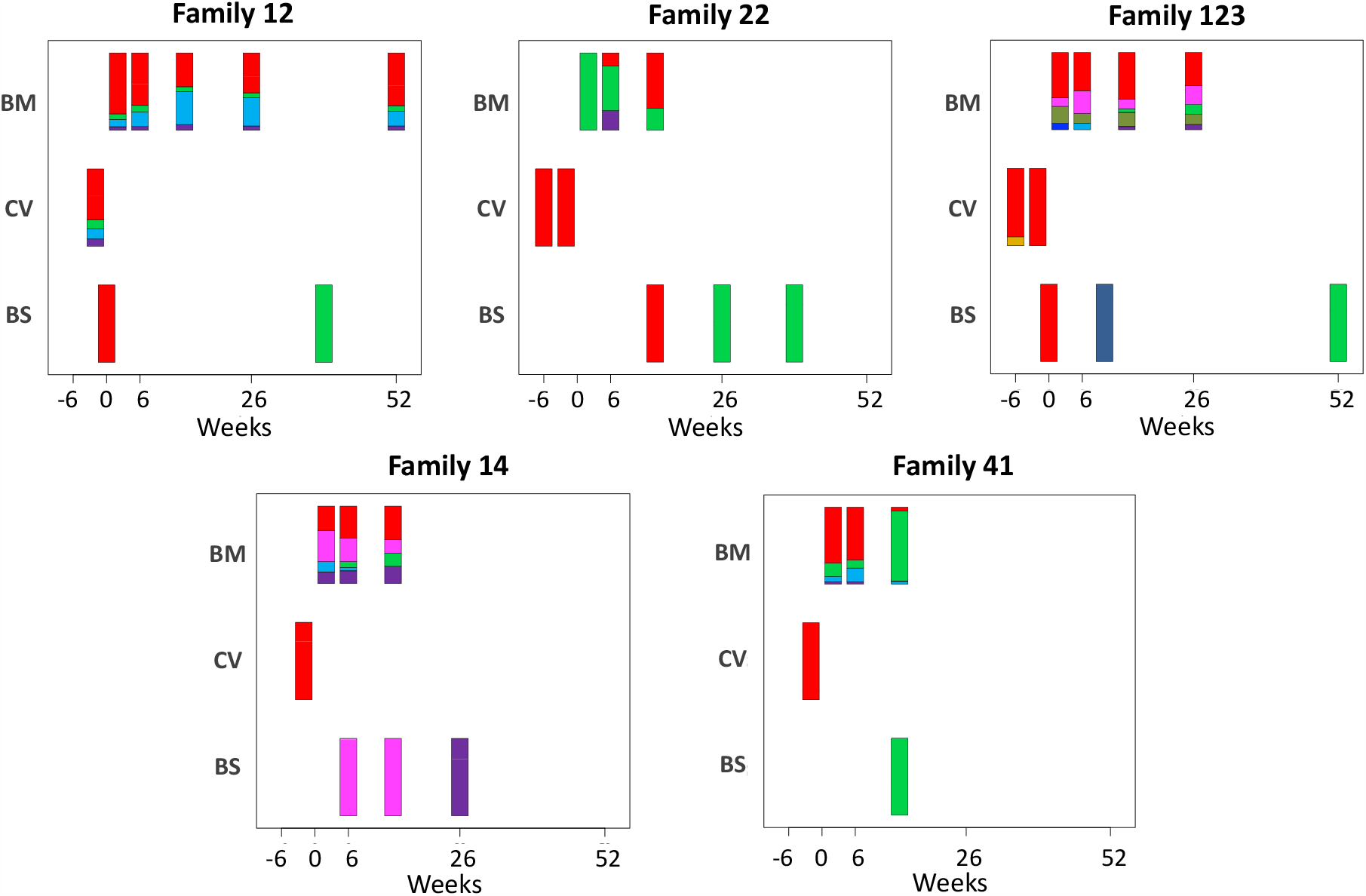
Abundance of haplotypes within each sample plotted for breast milk (BM), Cervix (CV) and Blood spots (BS). The timing of sampling is shown along the x axis. For ease of reference, the genotype containing the most abundant haplotype present in the cervix is coloured red for each family. Thereafter sequences that are genetically closest to the red genotype (Fig S5) are coloured magenta. Genotypes that are as distant from the cervical genotype as unrelated GenBank sequences are coloured shades of green, blue and purple. Single variants are coloured in shades of the nearest genotype.

### Transmission bottlenecks

CMV genomes from individual infant blood spots also showed lower diversity (Fig. S2), and predominance of one genotype (Fig. 4), including in samples with good sequence read depth e.g. Baby12 DEL and 9M, Baby14 6W,14W and 6M, Baby22 14W, Baby123 10W and 12M, (Table 1), indicating the likelihood of a bottleneck in mother-to-child transmission. Two infants (families 12 and 123 Fig. 1) who tested positive at birth were first infected with the genotype present in the greatest abundance in the cervix (Fig. 4 and Fig. S5). The same pattern was found in a third infant (family 22) whose first sample at two weeks of age tested positive (Figs. 2, 4 and S5). Interestingly, all three of these congenitally-infected infants were subsequently re-infected with distinct genotypes present in breast milk (Fig. 4). Two infants with initially two (family 14) and three (family 41) negative tests from birth onwards, first became positive at 6 and 10 weeks respectively. The genotypes detected in the blood spots from both of these infants were present in breast milk and differed from the most abundant genotype in cervix (Fig. 4).

### Subsampling to control for the impact of read depths

To determine the degree to which results were affected by the quality of sequence, we subsampled reads of different samples to show that sample diversity calculations are robust at read depths of ≥5 (Fig. S3); eight of the 18 blood spots and four of seven cervical samples had mean read depth ≥10 (Table 1) and all except one were of low diversity (Fig. S2). To determine the extent to which read depth affected haplotype frequencies, the 12-month breastmilk sample from mother 12, which had a mean read depth of 779.72 and five haplotypes (Fig. S5), was subsampled down to mean read depth <4 (Fig. S6). All of the haplotypes in this sample were present for read depths of 22 or more, with three haplotypes identified even at the lowest read depth. Nine out of ten cervical and blood spot samples from four families with read depths of >20 (Table 1), had either single genotypes or multiple closely related variants (Fig. 4) supporting previous conclusions around compartmentalization and transmission bottlenecks [26].

### Genotype compartmentalization

To look for evidence of inter-patient viral convergence by compartment, as has been observed previously [19], we used fixation index (FST) to compare the genetic similarity of individual genes of all subsets of two to five genotypes derived from different mother-baby pairs. P-values and false discovery rates for each pair were calculated using non-parametric bootstrapping. In order to compare various subsets, we computed a confidence weighted sum of FST (cwsFST) values for each subset. The distribution of cwsFST values is shown in Fig. S7. As can be seen, there are a large number of subsets with significant cwsFST values, far in excess of what is observed for scrambled sequences.

The sum weighted FST value for the subset of five genotypes that predominated in the cervical samples was not significantly different from other subsets, suggesting overall, that genotypes that predominated in the cervix of these women were not more closely related than genotypes found in breast milk. Intriguingly, however, the subset of cervical genotypes from mother-baby pairs 12, 22, and 123 had a sum weighted FST with a value greater than 99.6% of the other subsets, indicating a strong signal of inter-patient viral convergence. These genotypes were from the three mother-baby pairs with proven congenital infection based on first detection of CMV in the baby at ≤2 weeks of age, and in whom the baby’s genotype was identical to that predominating in cervix. In contrast, the predominant cervical genotypes from patients 41 and 14 showed low levels of relatedness with the first positive infant blood spot; these infant strains were more closely related to those from their mothers’ breast milk (Figs. 4 and S5).

The FST analysis identified 19 genes as likely to be contributing to the genetic similarity between congenitally transmitted genotypes from mothers 12, 22, 23 (FDR < 0.05) (Fig. 5). The comparison between these congenitally-transmitted and other genotypes generally yielded the same genes when the pairwise difference was varied to cluster haplotypes into more or fewer genotypes (Fig S8), suggesting that this finding is not an artefact of decisions about haplotype clustering.

**Fig. 5.**
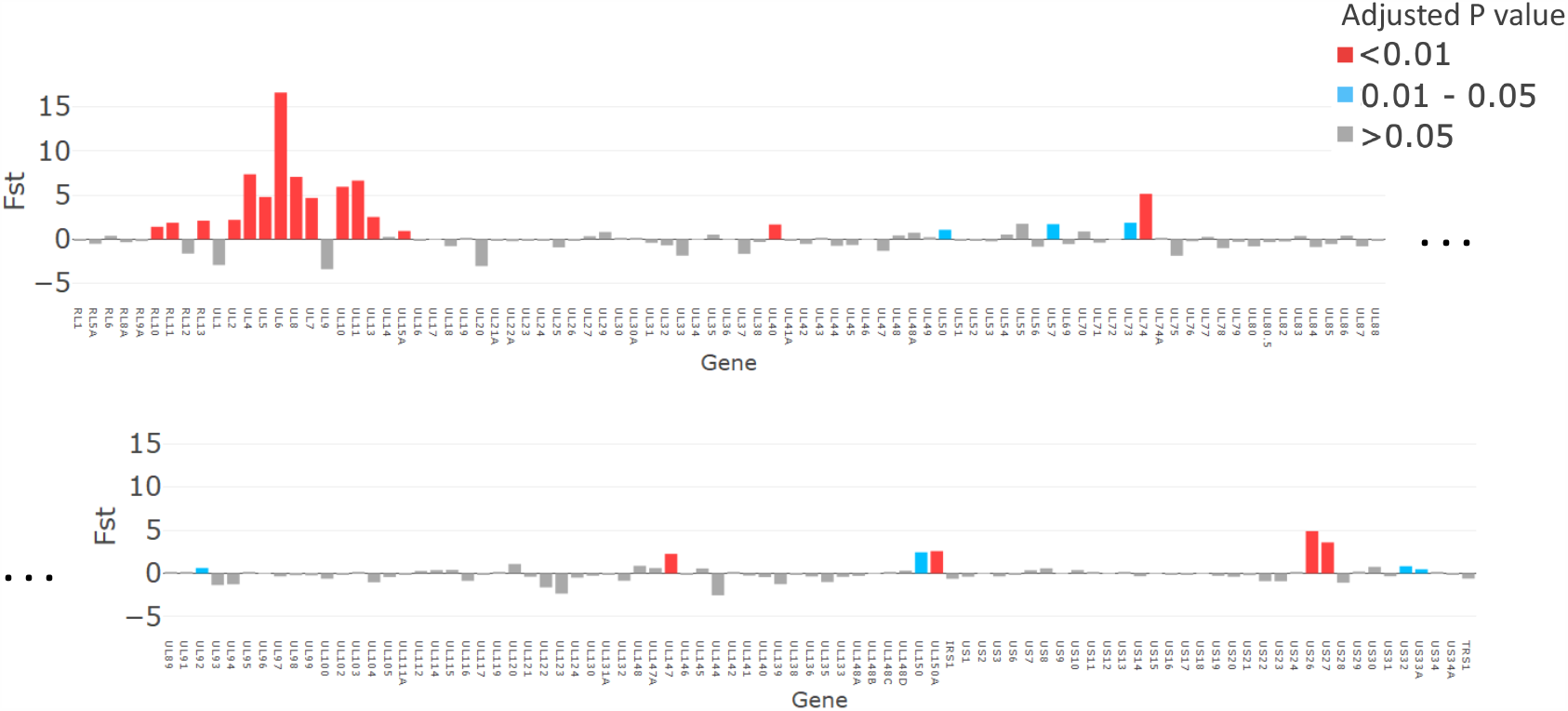
The magnitude of FST values plotted for each gene (x axis). P values, adjusted with false discovery rate are shown in Red for p <0.01, Grey for p >0.05 and turquoise for p=0.01-0.05.

## Discussion

We used next generation sequencing and haplotype reconstruction of individual CMV genomes, obtained from samples of HIV-infected women and their infants, to identify mixed infections, compartmentalization and distinct viral-genotype associations with transmission of CMV from mother-to-infant. Breast milk CMV showed high nucleotide diversity and, as has been previously reported [24], contained a mixture of viral genotypes, some of which were as genetically distant from each other as unrelated GenBank sequences and can therefore be considered distinct viral strains. Cervical samples were of low nucleotide diversity and dominated by a single viral genotype that was, with one exception, present in lower abundance in breast milk. Our data fit with most but not all [27] previous reports of CMV within-host compartmentalization based on genotyping of subgenomic fragments [28-31]. We found little evidence for widespread new superinfecting or reactivating viruses in these mothers. In line with findings from the immunosuppressed RhCMV monkey model of congenital infection, cCMVi [32] genotypes (strains) comprised families of closely related haplotypes. However, unlike the finding for congenitally transmitted gB and gL RhCMV variants, even where we found transmission of one genotype, maternal and infant haplotypes were not completely identical either in early, potentially congenital CMV infections, or in postnatally transmitted viruses from breastmilk. Neither were haplotypes sampled at different times from maternal breast milk conserved, suggesting a measure of de novo mutation in this patient group, in line with previous findings [19].

Our method of reconstructing viral haplotypes in serial samples provides insights into the natural history of CMV infection. While all mothers had mixtures of genotypes in breastmilk, the proportions changed over time for some (family 22 and 41) and remained more stable in others. Whether expanding genotypes in mothers 22 and 41 had been recently acquired is not known but would be consistent with incident reinfection. In contrast, all infants were initially infected with a single genotype (Fig. 4), supporting a bottleneck to CMV transmission [20, 32, 33]. Apparent reinfection by viruses present in breast milk occurred in all four infants with multiple samples (Fig. 4). We posit that the appearance of a new strain in an infant sampled from birth can confidently be interpreted as a newly-acquired exogenous virus rather than reactivation of a previously undetected one. In all cases, the reinfecting strains were genetically distant from and replaced the previously dominant strain (Fig. 4). Taken together with the rise and fall of infant CMV viral loads over time (Fig. 1), this pattern is consistent with immunity against the infants’ first CMV strain not being protective against reinfection with antigenically distinct strains, a concept that can be further tested. Of note, reinfection with the closely related strains also appears to occur readily with both human CMV and in animal models [15, 34]. Repeated reinfection with distinct strains may explain the high genetic variability observed between sequential samples in early sequencing studies of CMV genomes from congenitally-infected infants [18, 31].

Those infants who tested positive at <3 weeks from birth were congenitally-infected by definition[14]. In contrast, we cannot formally rule out cCMVi in the two others who were classified as having post-natal infection, since sensitivity of PCR detection of CMV DNA in newborn blood spots is only approximately 84% [35], and newborn saliva or urine were not available. However, this is unlikely given that only a small minority of infants have cCMVi, even among those born to HIV-infected women. Furthermore, it is striking that genotypes in babies with proven cCMVi were highly similar to maternal cervical genotypes, while those with negative tests for the first six weeks of life were not, and the strains detected later in the blood of these two infants were most similar to those in their mothers’ breast milk.

While it has previously been noted that a severe genetic bottleneck occurs during CMV transmission from mother to fetus or infant [19, 31, 36], it remains unknown whether CMV transmitted/founder virus populations share genotypic features that confer a fitness advantage for establishing an initial infection, such as seen in HIV [37]. Notwithstanding the apparent dominance of one genotype in each of the cervical samples, our analysis did not show evidence for inter-patient convergence of cervical genotypes per se. Rather the three cervical genotypes that were detected in babies 12, 22 and 123, who were infected at birth showed a higher level of genetic similarity than over 99.6% of other subset comparisons and much greater than would be expected by chance (black line) (Fig. S7). Nineteen genes (Fig. 5, Table 2) had particularly high (p<0.01) similarity scores. Twelve of the 19 genes with the highest similarity scores (Fig. 5) are part of the highly diverse RL11 gene family. Uniquely, RL11 genes form an island of linkage within the otherwise highly recombinant CMV genome [17]. Phylogeny of primate CMV RL11 complexes recapitulates the evolutionary history of the cognate host, suggesting it to be a potential driver of CMV co-evolution and speciation [17]. It is intriguing that RL11 family proteins influence tissue tropism [33] or are immunomodulatory [33, 38-42]. Together with its functional properties (Table 2) and extreme diversity [17], the possibility that within-species CMV RL11 variation may also influence within-host viral adaption to different compartments and/or transplacental transmission presents a tractable hypothesis that can now be tested. cCMVi is thought to occur primarily through maternal viremia followed by replication in placental cytotrophoblasts resulting in spread to the fetus [43]. We speculate that virus sampled in the cervix could be representative of CMV populations that are capable of infecting and crossing the placenta, rather than fetal infection ascending directly from the lower genital tract. Other genes with high similarity (FST) scores include US27, which codes for a G-protein-coupled receptor (GPCR) homologue that modulates signalling of the CXCR4 chemokine and may have a role during viral entry and egress [44], and US26 whose function is unknown. Less marked but still significantly different from non-congenitally transmitted strains, UL40 protein [45] modulates NK cell function. NK cells are the most abundant lymphocytes in placental tissue [43], while UL50 is also immunomodulatory [46, 47]. Finally, UL74, coding for glycoprotein O, which is highly significantly similar in all bar one comparisons (Fig. S8), is part of the gL/gH/g) complex which is critical for tropism and entry into both fibroblasts and epithelial cells [48]. Of interest, gB and gL which showed considerable diversity in the congenital RhCMV model were, as might be expected, not represented among the genes sharing significant genetic similarity in our analysis. One possibility that would unite our findings and those of the congenital RhCMV model is that CMV transmission bottlenecks are agnostic of variation in genes not implicated in transmission.

**Table 2.** Open Reading Frames (ORFs) identified by FST as being significantly more similar in strains transmitted prenatally. LD: Found to contain one of 33 hotspots of genetic linkage disequilibrium [17].

| ORF | LD | FUNCTION |
| --- | --- | --- |
| UL10 | Y | Putative membrane glycoprotein, Immunosuppressive impairs T cell function [41] |
| UL11 | Y | Membrane glycoprotein modulation of T cell signalling/function [42, 49] |
| UL13 |  | Unknown function |
| UL4 | Y | Putative membrane glycoprotein [39] |
| UL5 |  | Putative membrane glycoprotein [39] |
| UL6 | Y | Putative membrane glycoprotein [39] |
| UL7 | Y | Membrane glycoprotein, modulates chemo-and/or cytokine signalling function [40] |
| UL8 | Y | Transmembrane glycoprotein. Inhibits proinflammatory cytokines [40] |
| US26 |  | Unknown function |
| US27 | Y | Membrane glycoprotein Activates CXCR4 signalling to increase HCMV replication [44] |
| UL150A |  | Fibroblast and Epithelial cell entry [50] |
| UL2 |  | Putative membrane glycoprotein [39] |
| RL11 | Y | Membrane glycoprotein. Binds IgG Fc domain involved in immune regulation [39] |
| UL147 | | α-chemokine homologue [51, 52] |
| UL40 |  | Control of NK recognition [45] |
| RL13 | Y | Glycoprotein, repression of replication, bind IgG domain immune regulation [33, 38] |
| RL10 |  | Membrane glycoprotein |
| UL57 |  | Ss DNA binding protein [39] |
| UL50 |  | Nuclear Egress complex. Reduces interferon mediated antiviral effect [47] |

Being born to an HIV-infected women is a major risk factor for cCMVi as well as long term CMV-related complications, whether or not the child acquires HIV [8, 9]. We show here that, irrespective of the route of first infection, HIV-exposed uninfected (HEU) children frequently acquire repeated infections with different CMV viruses within the first year of life. Preliminary evidence suggests that breast milk of HIV-uninfected women may have lower CMV viral loads and carry fewer strains [53]. If this is true, the possibility that HEU, as well as HIV-infected, infants are exposed to greater numbers of CMV strains during infancy as compared with HIV-uninfected infants, may provide an explanation for their worse clinical outcomes, a hypothesis that can now be tested in prospective studies. Similarly, these methods promise to be invaluable for studying the role of maternal CMV reinfection during pregnancy, a question of central importance in the field [12].

This study potentially provides several new insights into the pathogenesis of CMV infection. However, the study is limited by the small number of subjects, the fact that all women were HIV-1 infected and the lack of samples and data to absolutely confirm the route of CMV acquisition by these infants. Because we were only able to analyse maternal breast milk, cervical samples and infant blood, and only intermittently, it is possible that some transmitted viral variants were not captured. Some, particularly cervical and blood spot samples, had low CMV viral loads and, as a result, suboptimal genome coverage. Mapping data confirmed that in these cases sequence loss was random, excluding the possibility of systematic bias. To further address this potential bias, we subsampled samples with good coverage to identify read-depth thresholds above which the diversity estimation is robust and haplotype frequency to 5% and above is preserved (Supplementary Figs. 3 and 7). Analysis of only those samples with read depths above the identified thresholds supported our overall conclusions. The quality of the sequence and the numbers of samples allowed for conclusions to be drawn at gene level only and precluded robust identification of putative motifs or single nucleotide polymorphisms associated with biological differences.

In summary, by reconstructing the individual CMV haplotypes we found evidence for mixed CMV infection in HIV-infected women, and compartmentalization of viral strains between cervical and breast milk. Infants appeared usually to acquire one virus genotype initially, indicating a transmission bottleneck, though subsequent reinfection with a second virus from maternal breast milk was common. We also found that viruses transmitted congenitally resembled the virus genotypes that were present at highest abundance in cervix, and shared genetic features that distinguished them from CMV strains predominating in breast milk and in the cervices of women whose infants were apparently first infected post-partum. These data provide new testable insights into the pathogenesis of CMV transmission from mothers to their infants, as well as tools to unravel the importance of viral diversity for reinfection and congenital transmission, questions that are central to the development of a vaccine to prevent the global burden of disease due to CMV.

## Materials and Methods

Samples were approved for research by the Institutional Review Board of the University of Washington and the Ethics and Research Committee of Kenyatta National Hospital IRB NCT00530777 and sequenced under the ULCP Biobank REC approval. Approval for use of anonymised residual diagnostic specimens were obtained through the University College London/University College London Hospitals (UCL/UCLH) Pathogen Biobank National Research Ethics Service Committee London Fulham (Research Ethics Committee reference: 12/LO/1089). Informed patient consent was not required.

### Patient specimens

Mother-child pairs were selected from a randomized, placebo-controlled trial to determine the impact of twice-daily valacyclovir (500 mg) on breast milk HIV RNA viral load in HIV-1/HSV-2 co-infected women (NCT 00530777). Trial design, participant characteristics, and follow-up have been reported elsewhere, [21-23] and the University of Washington Institutional Review Board and Kenyatta National Hospital Research and Ethics Committee approved the research. Women received short course antiretrovirals for prevention of mother-to-child HIV transmission, but no women or infants received combination antiretroviral therapy, as the study was conducted before recommendations for universal treatment. All women were HIV-1, HSV-2 and CMV co-infected. For this CMV genomics study, we selected 5 mother-infant pairs from the placebo arm with HIV-exposed uninfected infants, who had well-defined timing of infant CMV infection. Women had cervical swabs and blood specimens collected at 34- and 38-weeks gestation. Maternal blood and infant dried blood spots were collected delivery, then postpartum at 2, 6, 10, 14, 24, 36, and 52 weeks. Breast milk was collected at all times after delivery. Blood plasma, cervical swabs, and breast milk supernatant (whey) was cryopreserved at –80 C for the study of HIV and other co-infections.

### DNA extraction and CMV DNA measurement

Viral nucleic acids were extracted from blood plasma, dried blood spots, breast milk supernatant and cervical swabs as previously described using the Qiagen UltraSens Viral Nucleic Acid extraction kit [22]. Quantitative real-time PCR was used to measure CMV DNA levels in these specimens [22].

### Sure-select sequencing

Hybridization and library preparation was performed as previously described [54]. Briefly, extracted DNA was sheared by acoustic sonication (Covaris e220, Covaris Inc.). DNA fragments underwent end-repair, A’-tailing, and (Illumina) adaptor ligation. DNA libraries were hybridised with biotinylated 120-mer custom RNA baits designed using all available CMV full genomes in Genbank for 16-24 hrs at 65°C and subsequently bound to MyOne™ Streptavidin T1 Dynabeads™ (ThermoFisher Scientific). Following washing, libraries were amplified (18 cycles) to generate sufficient input material for Illumina sequencing. Paired end sequencing was performed on an Illumina MiSeq using the 500 cycle v2 Reagent Kit (Illumina, MS-102-2003). Samples were sequenced in four different batches by family group.

Reads generated were quality checked and mapped to the Merlin Reference sequence followed by removal of duplicates using the CLC Genomics Workbench ver. 10.1. Consensus sequence was extracted with a minimum coverage of 2X. Consensus sequences along with other Genbank reference sequences were aligned using MAFFT 7.212 [55] and refined by manual editing.

### Clustering

Pairwise distances between sequences were calculated using the dist.dna function from R package Ape v.5.3 [56]. Sequences were clustered using multidimensional scaling as implemented by the cmdscale function from R package Stats v.3.6 [57].

### Nucleotide diversity

Nucleotide diversity was calculated by fitting the observed variant frequency spectrum to the mixture of two distributions, one representing sequencing errors (represented by a Beta distribution), the other representing true diversity (represented by a four-dimensional Dirichlet distribution plus delta function, the latter representing invariant sites). The parameters for these two distributions were optimized by maximizing the log likelihood. This framework allows all of the sequencing data to be used and does not require pre-filtering the data to remove sites with low read depth or few variants resulting in the favorable robustness to read depth, as shown in Fig. S3. Software is available for download at GitHub Repository, https://github.com/ucl-pathgenomics/NucleotideDiversity.

### Haplotype reconstruction

Haplotype reconstruction was accomplished using HaROLD with default settings [25]. Details of this procedure are described in the associated publications. In brief, HaROLD employs a two-step process. The first step is based on the assumption that there are a limited number of haplotypes that are the same for all of the samples from a given mother/ child data set, so that the differences in the frequencies of polymorphisms represent different mixtures of these haplotypes. By taking advantage of the co-variation of variant frequencies, HaROLD creates a set of haplotypes for each of the data sets, optimized so that linear combinations of these haplotypes can best account for the observed variant frequencies. The number of haplotypes is chosen to maximize the log likelihood of the observed frequencies. The second step involves relaxing the assumption of constant haplotypes, with each sample treated individually. For each sample, reads are assigned probabilistically to the various haplotypes generated by the first step. These haplotype sequences and frequencies are then adjusted based on the assigned reads. The reads are then re-assigned to these adjusted haplotypes, and the procedure is repeated until convergence. During this process, haplotypes can be merged if that decreases the Akaike Information Criterion (AIC) [58]. This procedure results in a set of haplotypes for each sample, loosely based on the haplotypes derived from the first step.

### Haplotype trees

Maximum Likelihood trees of the haplotypes from each family were computed using RaxML v8.2.10, implementing the GTR model, with 1000 bootstrap replicates [59].

### Haplotype clustering

The haplotypes for each mother/baby data set were divided into genotypes. We calculated the pairwise evolutionary distance (the sum of distances on the evolutionary tree between the haplotypes and their latest common ancestor) for all pairs of haplotypes in each family. As shown in Fig. S9, the observed distribution of such pairwise distances fits the sum of a Gamma distribution (69.3%, alpha = 19.5, beta = 0.0015) and an exponential distribution (30.7%, mean = 0.01), indicative of two classes of relationships – pairs of sequences that are highly similar, modelled by the exponential, representing small accumulated variations, and pairs that are more distinct, represented by the Gamma distribution. We chose the crossing point of these two distributions, at a cut-off distance of 0.017, as differentiating small variations from larger differences (Fig. S9). We then grouped the haplotypes into clusters so that all members of a cluster have a pairwise evolutionary distance with all other members less than 0.017, resulting in 26 clusters which we refer to as genotypes. We used these groups to assign colours to the different haplotype-clusters (genotypes) in Fig. 4 and Fig. S5.

We used FST to identify sequence characteristics associated with sets of genotypes. Consensus sequences were constructed for each genotype. FST values, representing the genetic difference between a subset of genotypes and the other genotypes, were calculated for each gene. P-values and corresponding false discovery rates were estimated by non-parametric bootstrapping, through scrambling the bases at each position amongst the clusters. The results are shown for the 26 genotypes obtained with a cut-off distance of 0.017; changing this cut-off resulted in increased or decreased numbers of genotypes, but yielded similar results, especially for the more confident identifications (Fig. S8).

### Evaluating the similarity between subsets of genotypes

We use FST values to identify similarities between individual genes from subsets of genotypes compared with the other genotypes. In order to compare the magnitude of the similarities of different subsets, we would like to take the sum of the FST values for all genes where the similarities are real and not the result of random associations. As we cannot definitively identify these genes, we instead consider the sum of the FST values for all genes weighted by our confidence that the FST value is significant, represented as one minus the false discovery rate.

## Data Availability

Sequence reads have been deposited in NCBI Sequence Read Archive under BioProject ID PRJNA605798.

## Acknowledgments

We acknowledge the support of the MRC/NIHR UCLH/UCL Biomedical Research Centre funded Pathogen Genomics Unit. This work was funded by EUFP7 grant 304875 (PI Breuer), Wellcome Trust grant 204870 (PI Griffiths), NIH National Institute of Allergy and Infectious Diseases grant AI087369 (PI Slyker), AI027757 (PI Slyker, Holmes), AI076105 and K24 AI087399 (Farquhar), National Institute of Child Health and Human Development HD057773–01, HD054314 (Farquhar). JP is funded by a Rosetrees Trust PhD Studentship M876. SM and J Bryant are funded by Henry Wellcome fellowships. J Breuer receives funding from the UCL/UCLH NIHR Biomedical Research Centre.

## Data availability

Sequence reads have been deposited in NCBI Sequence Read Archive under BioProject ID PRJNA605798.

All software used are available for download at GitHub Repository, https://github.com/ucl-pathgenomics/NucleotideDiversity and https://github.com/ucl-pathgenomics/HAROLD.

---

**Fig. S1.**
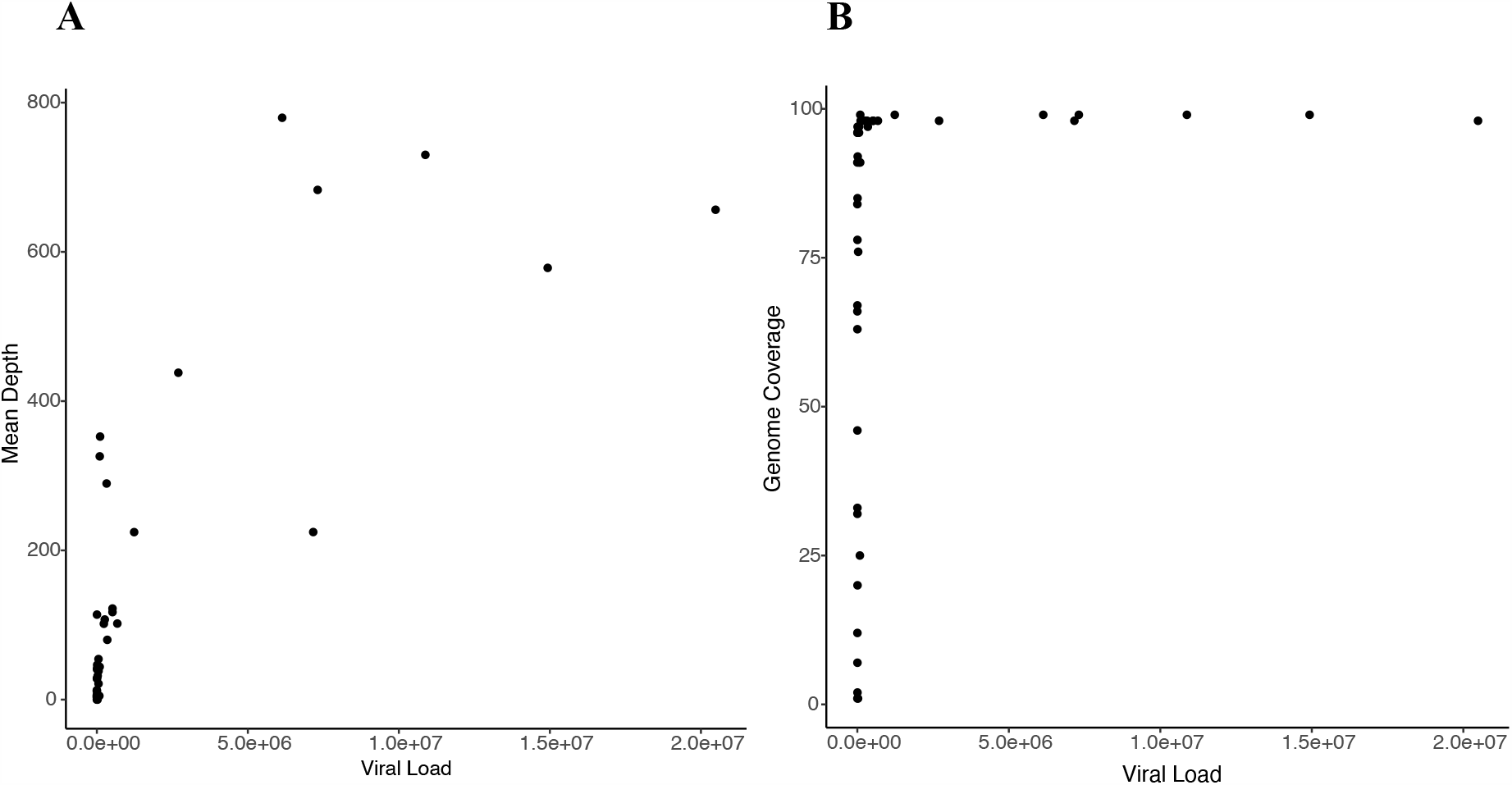
Scatter plots showing relationship between input viral load and (**A**) mean read depth and **(B**) genome coverage respectively.

**Fig. S2.**
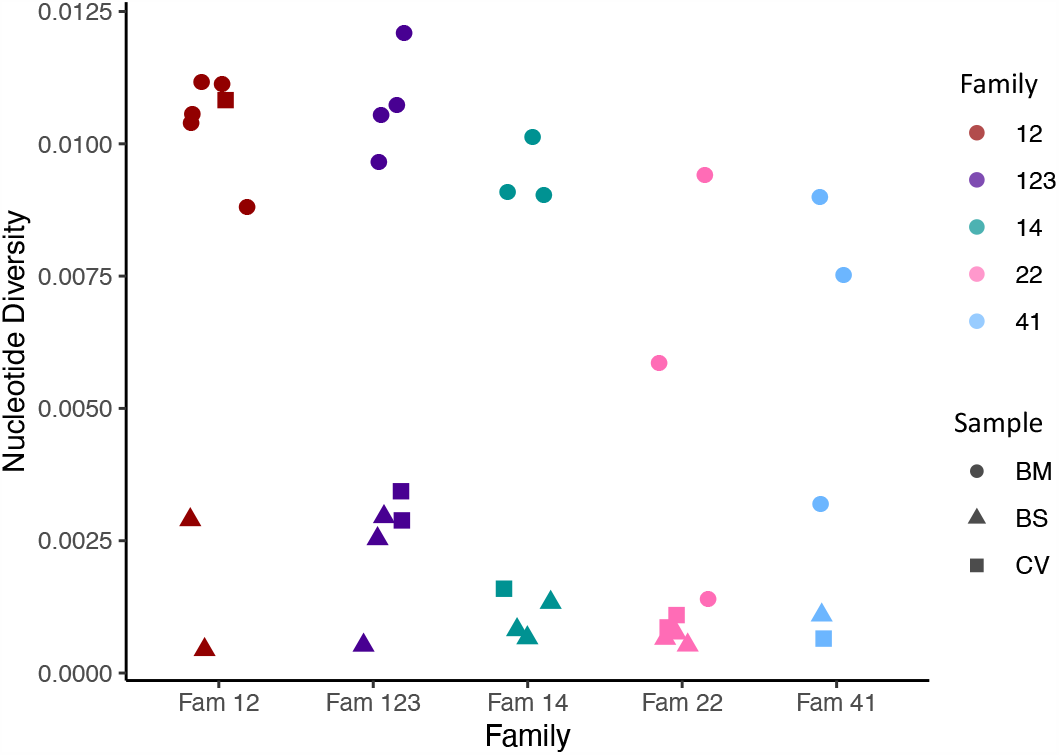
Within sample nucleotide diversity shown by family (colour) and sample type (symbol). BM; breast milk, CV; cervix, BS; baby blood spot. The figure shows that most cervical and blood spot samples are of low diversity, while most breast milk samples are of high diversity

**Fig. S3.**
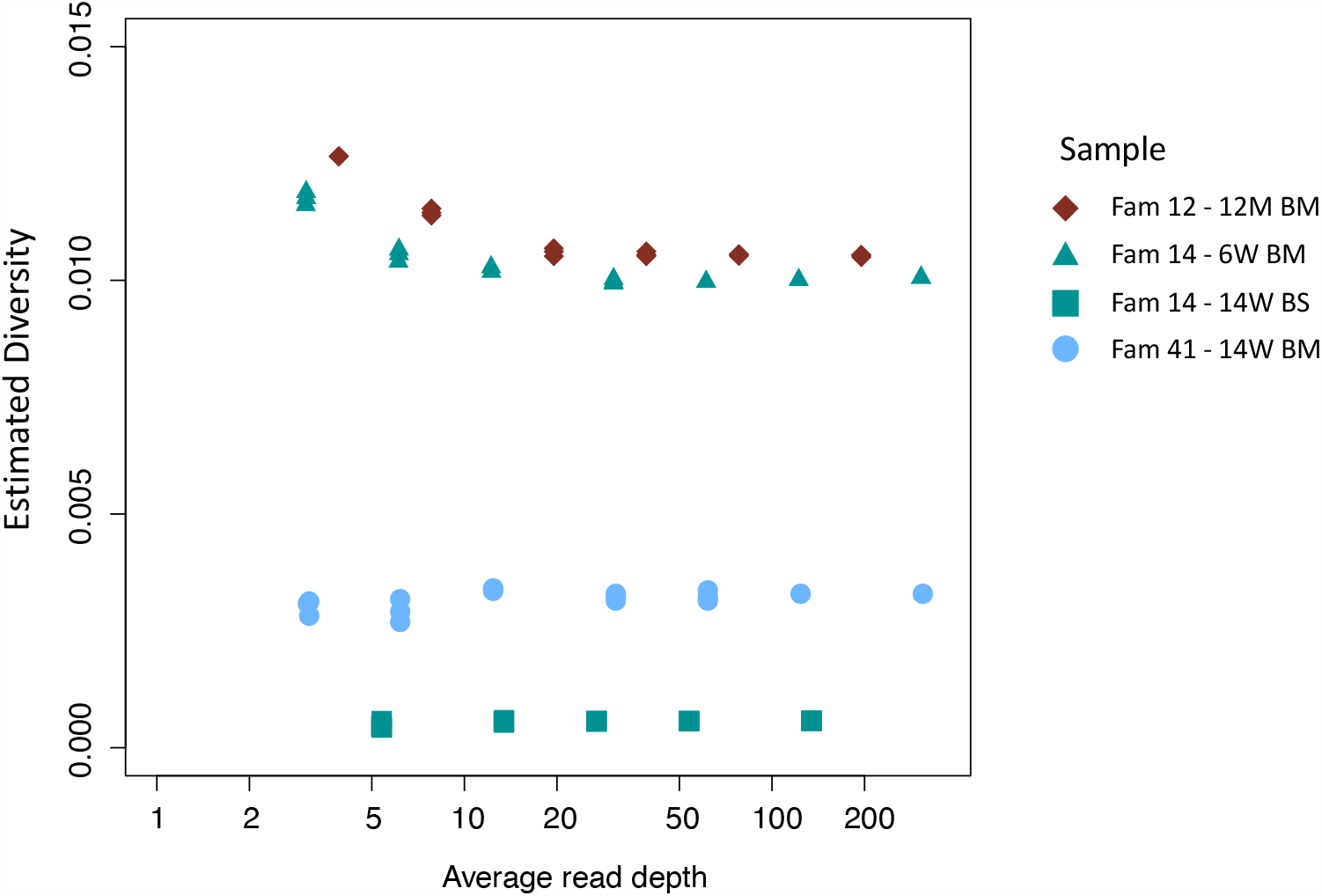
Effect of down-sampling on estimated diversity. Samples tested include family 14 14W BS (green squares), family 41 14W BM (blue dots), family 14 6W BM (green triangles), family 12 12M BM (maroon diamonds) all of which had initial read depths of 150 or more. The estimated diversity is relatively insensitive to read depth; in particular, down-sampling of high read-depth samples shows no tendency of the analysis to underestimate the diversity of low read-depth samples. This indicates that the low diversity observed in many of the CV and BS samples is not an artefact but is rather consistent with the presence of significant bottlenecks.

**Fig. S4.**
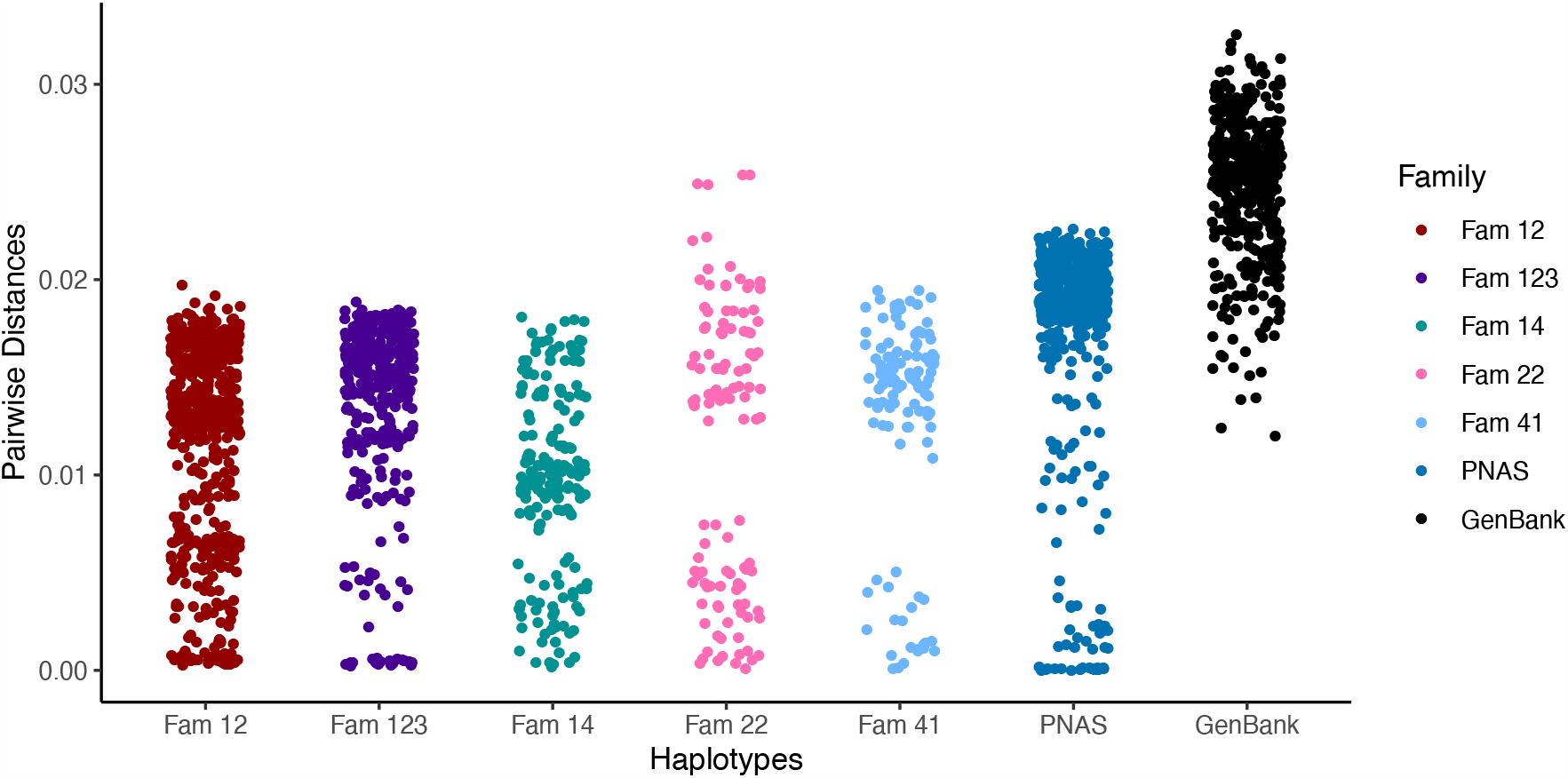
Pairwise differences between haplotypes within a family. Distances are compared with random GenBank sequences and sequences previously analyzed by the same pipeline and reported [20]. Higher values are similar to those seen between unrelated database sequences and indicate the presence of distinct strains.

**Fig. S5.**
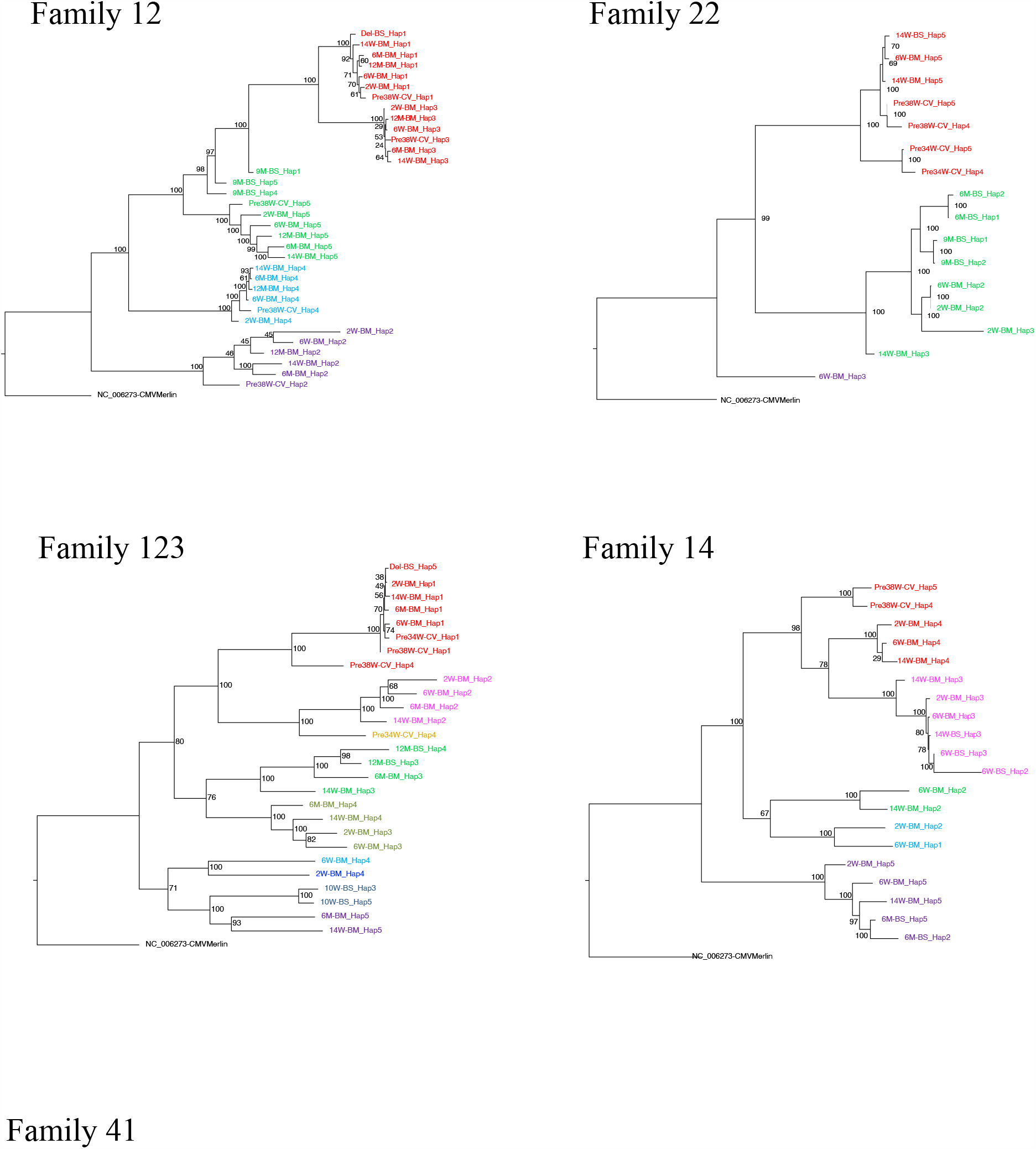
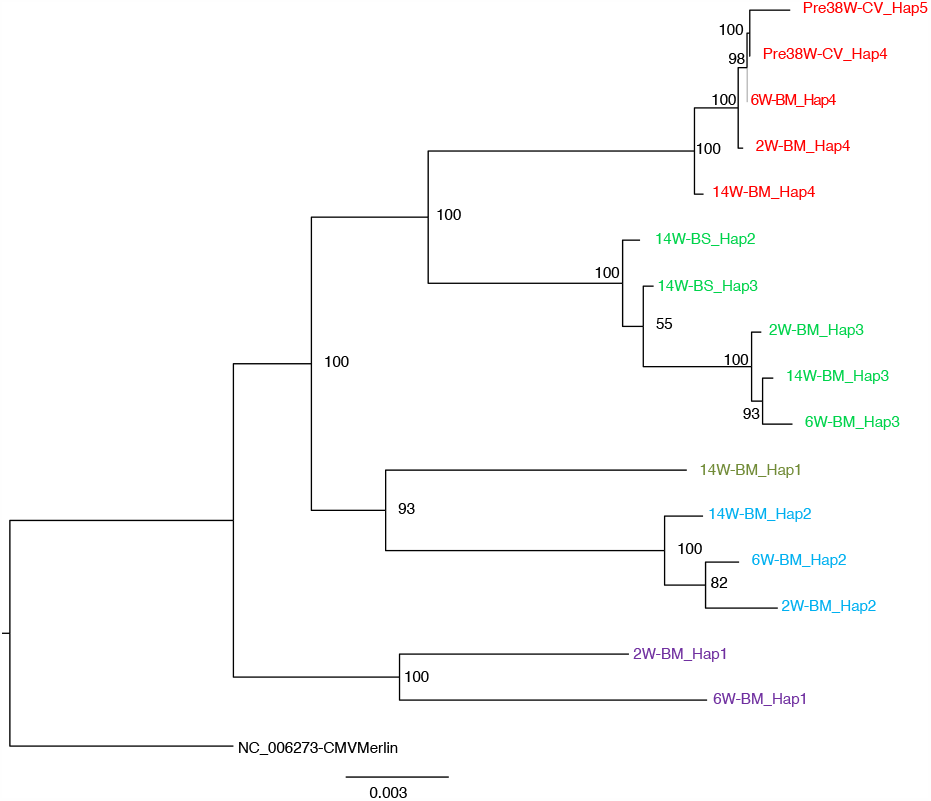
Maximum-likelihood phylogenetic tree to show haplotypes clusters (genotypes). By convention, the genotype most prevalent in cervix was colored red for each family. Genotypes were designated where a distinct cluster of related haplotypes (pairwise distance ≤0.017) occurred with a bootstrap value of 100 (see methods and supplementary figure 8). The genotype containing the most abundant haplotype present in the cervix is coloured red for each family. Thereafter sequences that are genetically closest to the red genotype are coloured magenta. Genotypes that are as distant from the cervical genotype as unrelated GenBank sequences are coloured shades of green, blue and purple. The number of clusters between 18 and 34 did not affect subsequent conclusions about genetic similarly between cervical versus other strains (see Fig. S8).

**Fig. S6.**
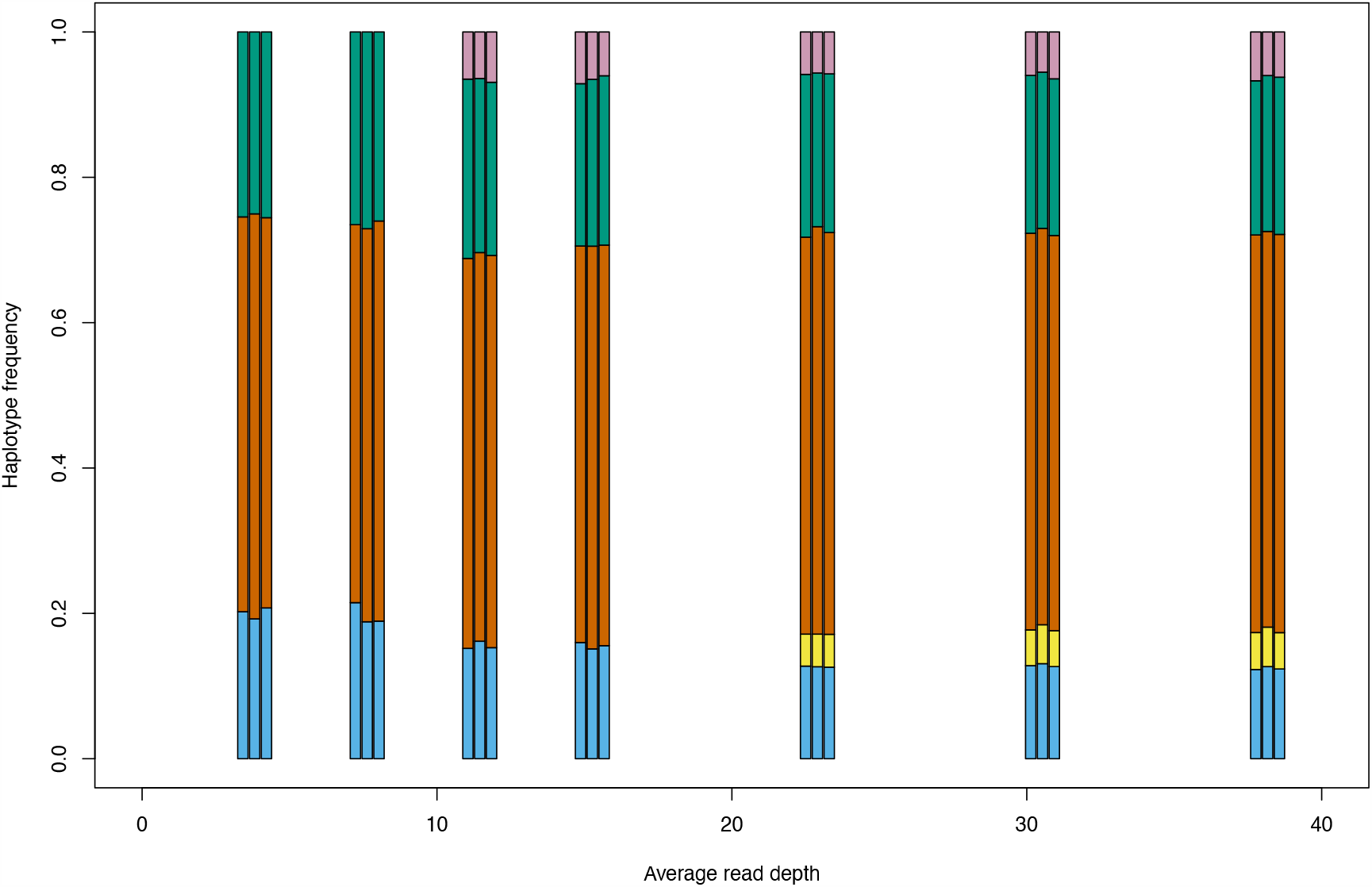
Boxplot showing number of haplotypes reconstructed in relation to read depth. Analysis was performed on the 12-month breastmilk sample from family 12.

**Fig. S7:**
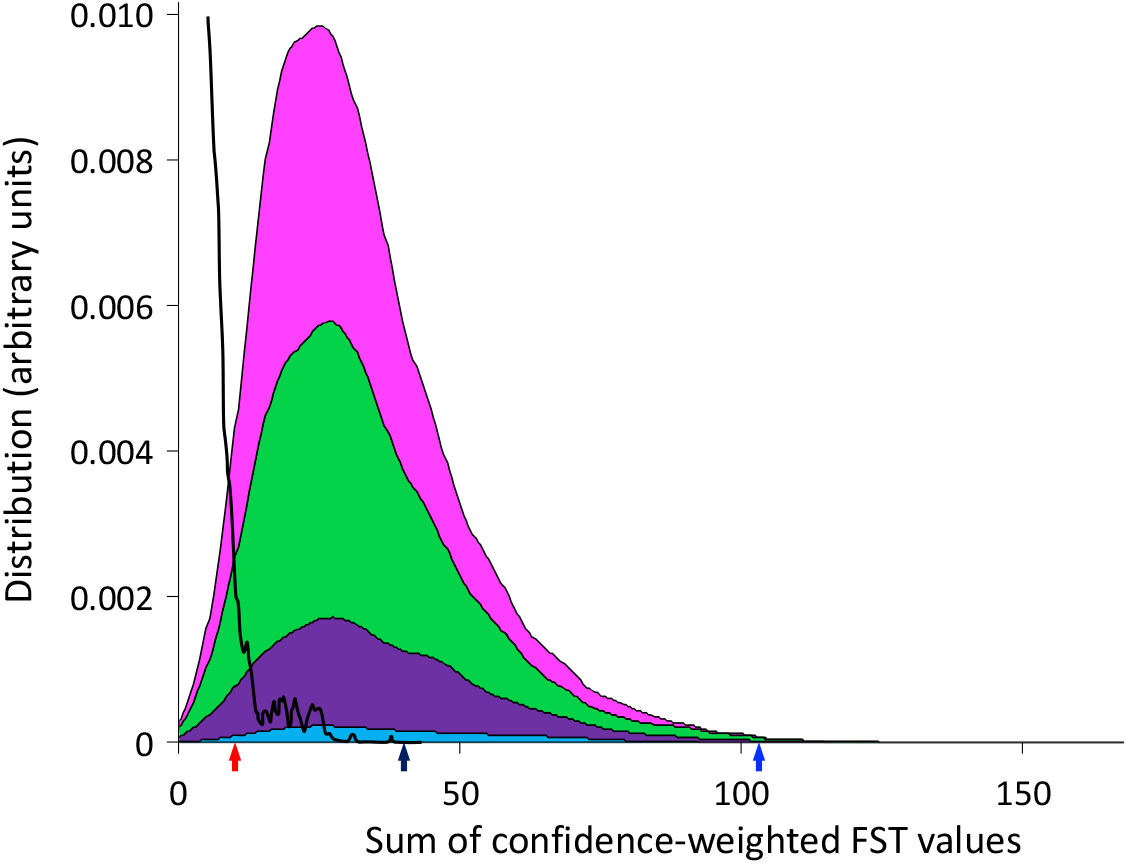
Distribution of confidence-weighted sums of FST (cwsFST) values for all subsets of two (cyan), three (purple), four (green) and five (magenta) genotypes from different mother-baby pairs. For comparison, we also show the distribution obtained when the genotype sequences corresponding to each mother-baby pair are scrambled (black line). Arrows mark the values for the five genotypes that predominated in the cervical samples (black), the three predominant genotypes from cervical samples for mother-baby pairs 12, 22, and 123 (blue), and the two predominant genotypes from cervical samples for mother-baby pairs 14 and 41 (red).

**Fig S8.**
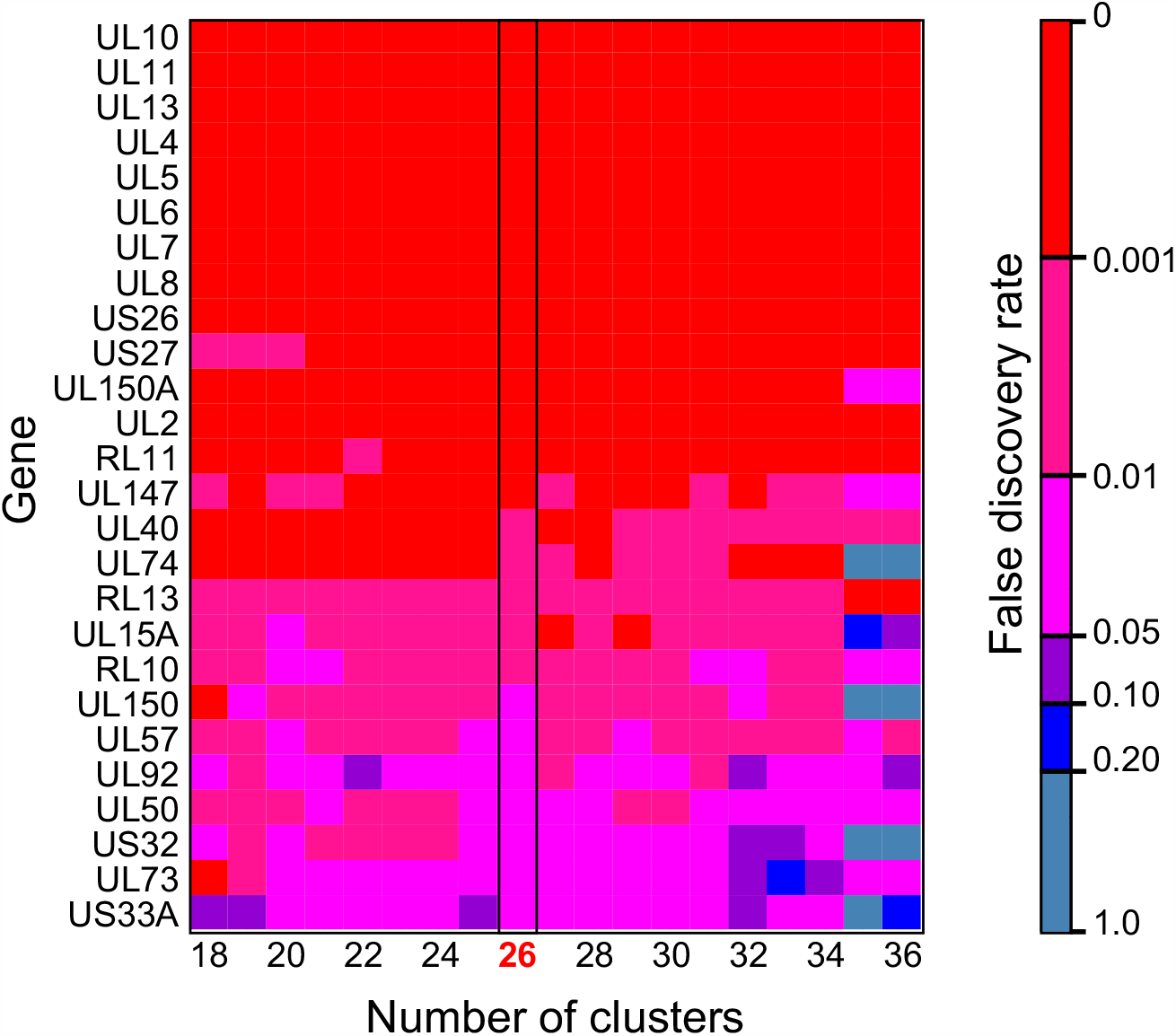
Heatmap showing genes identified as significant in F_ST_ analysis are robust to changes in the number of clusters. Colors indicated the false discovery rate value, red = <0.001; magenta = 0.001-0.01; pink = 0.01-0.05; purple = 0.05-0.1; blue = 0.1-0.2; grey = >0.2.

**Fig. S9.**
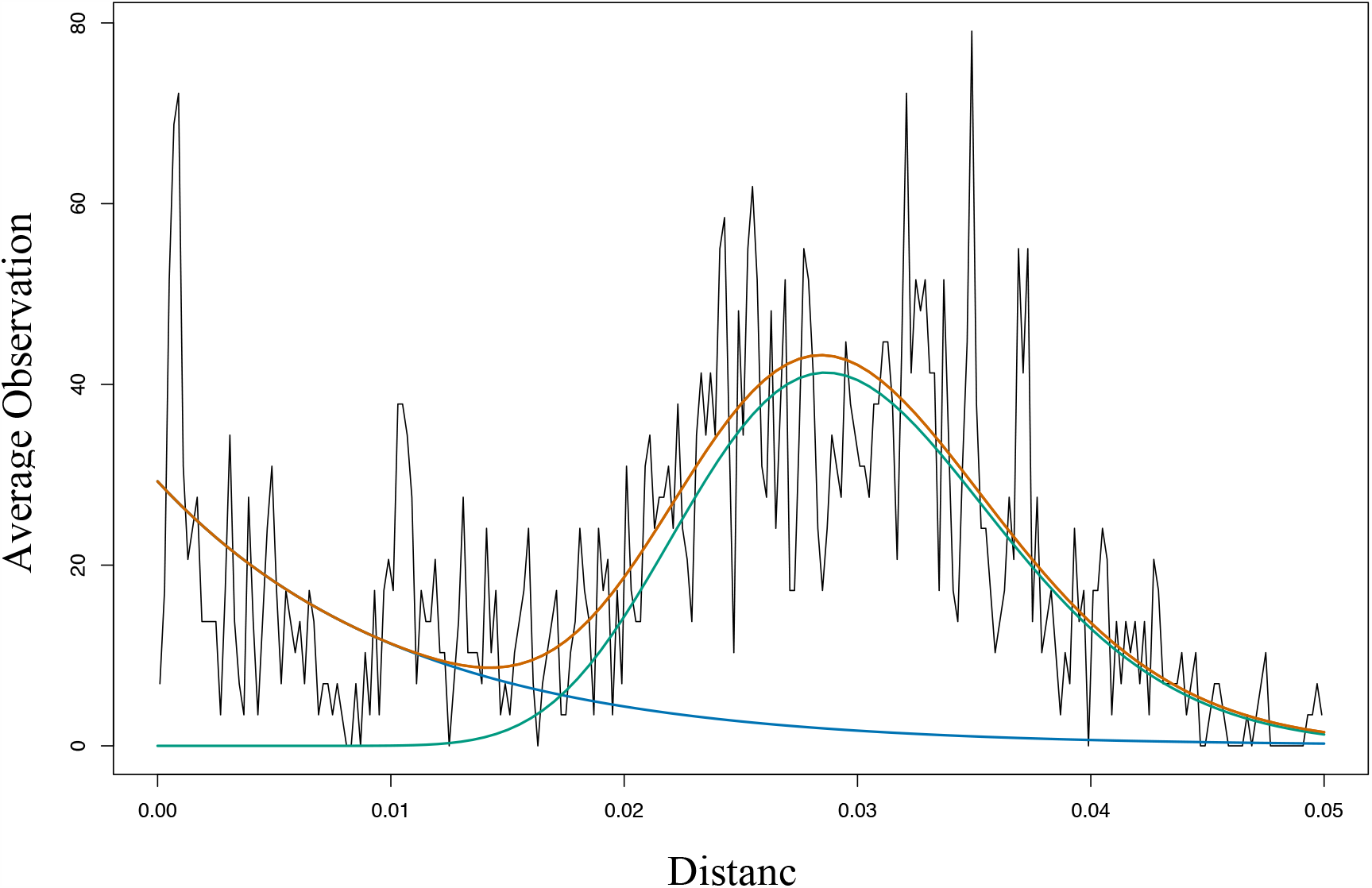
Distribution of pairwise evolutionary distances for haplotypes within families. Black, observed distribution of pairwise evolutionary distances; green, gamma distribution; blue, exponential distribution; orange, sum of Gamma distribution plus Exponential Distribution.

## References

1. Morton CC, Nance WE. Newborn hearing screening--a silent revolution. N Engl J Med. 2006;354(20):2151–64. Epub 2006/05/19. doi: 10.1056/NEJMra050700. PubMed PMID: 16707752.

2. Boppana SB, Ross SA, Fowler KB. Congenital cytomegalovirus infection: clinical outcome. Clinical infectious diseases : an official publication of the Infectious Diseases Society of America. 2013;57 Suppl 4:S178–81. Epub 2013/12/07. doi: 10.1093/cid/cit629. PubMed PMID: 24257422.

3. Dollard SC, Grosse SD, Ross DS. New estimates of the prevalence of neurological and sensory sequelae and mortality associated with congenital cytomegalovirus infection. Rev Med Virol. 2007;17(5):355–63. Epub 2007/06/02. doi: 10.1002/rmv.544. PubMed PMID: 17542052.

4. Gantt S, Orem J, Krantz EM, Morrow RA, Selke S, Huang ML, et al. Prospective Characterization of the Risk Factors for Transmission and Symptoms of Primary Human Herpesvirus Infections Among Ugandan Infants. J Infect Dis. 2016;214(1):36–44. doi: 10.1093/infdis/jiw076. PubMed PMID: 26917575; PubMed Central PMCID: PMCPMC4907408.

5. Gantt S, Leister E, Jacobsen DL, Boucoiran I, Huang ML, Jerome KR, et al. Risk of congenital cytomegalovirus infection among HIV-exposed uninfected infants is not decreased by maternal nelfinavir use during pregnancy. J Med Virol. 2016;88(6):1051–8. doi: 10.1002/jmv.24420. PubMed PMID: 26519647; PubMed Central PMCID: PMCPMC4818099.

6. Slyker JA, Richardson B, Chung MH, Atkinson C, Asbjornsdottir KH, Lehman DA, et al. Maternal Highly Active Antiretroviral Therapy Reduces Vertical Cytomegalovirus Transmission But Does Not Reduce Breast Milk Cytomegalovirus Levels. AIDS Res Hum Retroviruses. 2017;33(4):332–8. Epub 2016/11/01. doi: 10.1089/AID.2016.0121. PubMed PMID: 27796131; PubMed Central PMCID: PMCPMC5372773.

7. Richardson BA, John-Stewart G, Atkinson C, Nduati R, Asbjornsdottir K, Boeckh M, et al. Vertical Cytomegalovirus Transmission From HIV-Infected Women Randomized to Formula-Feed or Breastfeed Their Infants. J Infect Dis. 2016;213(6):992–8. doi: 10.1093/infdis/jiv515. PubMed PMID: 26518046; PubMed Central PMCID: PMCPMC4760415.

8. Garcia-Knight MA, Nduati E, Hassan AS, Nkumama I, Etyang TJ, Hajj NJ, et al. Cytomegalovirus viraemia is associated with poor growth and T-cell activation with an increased burden in HIV-exposed uninfected infants. AIDS. 2017;31(13):1809–18. Epub 2017/06/14. doi: 10.1097/QAD.0000000000001568. PubMed PMID: 28609400; PubMed Central PMCID: PMCPMC5538302.

9. Gompels UA, Larke N, Sanz-Ramos M, Bates M, Musonda K, Manno D, et al. Human cytomegalovirus infant infection adversely affects growth and development in maternally HIV-exposed and unexposed infants in Zambia. Clinical infectious diseases : an official publication of the Infectious Diseases Society of America. 2012;54(3):434–42. doi: 10.1093/cid/cir837. PubMed PMID: 22247303; PubMed Central PMCID: PMC3258277.

10. Hsiao NY, Zampoli M, Morrow B, Zar HJ, Hardie D. Cytomegalovirus viraemia in HIV exposed and infected infants: prevalence and clinical utility for diagnosing CMV pneumonia. J Clin Virol. 2013;58(1):74–8. Epub 2013/06/04. doi: 10.1016/j.jcv.2013.05.002. PubMed PMID: 23727304.

11. Kenneson A, Cannon MJ. Review and meta-analysis of the epidemiology of congenital cytomegalovirus (CMV) infection. Rev Med Virol. 2007;17(4):253–76. Epub 2007/06/21. doi: 10.1002/rmv.535. PubMed PMID: 17579921.

12. Britt WJ. Congenital Human Cytomegalovirus Infection and the Enigma of Maternal Immunity. J Virol. 2017;91(15). doi: 10.1128/JVI.02392-16. PubMed PMID: 28490582; PubMed Central PMCID: PMCPMC5512250.

13. de Vries JJ, van Zwet EW, Dekker FW, Kroes AC, Verkerk PH, Vossen AC. The apparent paradox of maternal seropositivity as a risk factor for congenital cytomegalovirus infection: a population-based prediction model. Rev Med Virol. 2013;23(4):241–9. doi: 10.1002/rmv.1744. PubMed PMID: 23559569.

14. Boppana SB, Fowler KB, Britt WJ, Stagno S, Pass RF. Symptomatic congenital cytomegalovirus infection in infants born to mothers with preexisting immunity to cytomegalovirus. Pediatrics. 1999;104(1 Pt 1):55–60. PubMed PMID: 10390260.

15. Boucoiran I, Mayer BT, Krantz EM, Marchant A, Pati S, Boppana S, et al. Nonprimary Maternal Cytomegalovirus Infection After Viral Shedding in Infants. Pediatr Infect Dis J. 2018;37(7):627–31. Epub 2018/06/12. doi: 10.1097/INF.0000000000001877. PubMed PMID: 29889809; PubMed Central PMCID: PMCPMC6016842.

16. Barbosa NG, Yamamoto AY, Duarte G, Aragon DC, Fowler KB, Boppana S, et al. Cytomegalovirus Shedding in Seropositive Pregnant Women From a High-Seroprevalence Population: The Brazilian Cytomegalovirus Hearing and Maternal Secondary Infection Study. Clinical infectious diseases : an official publication of the Infectious Diseases Society of America. 2018;67(5):743–50. Epub 2018/03/01. doi: 10.1093/cid/ciy166. PubMed PMID: 29490030; PubMed Central PMCID: PMCPMC6094000.

17. Lassalle F, Depledge DP, Reeves MB, Brown AC, Christiansen MT, Tutill HJ, et al. Islands of linkage in an ocean of pervasive recombination reveals two-speed evolution of human cytomegalovirus genomes. Virus Evol. 2016;2(1):vew017. Epub 2016/06/15. doi: 10.1093/ve/vew017. PubMed PMID: 30288299; PubMed Central PMCID: PMCPMC6167919.

18. Pokalyuk C, Renzette N, Irwin KK, Pfeifer SP, Gibson L, Britt WJ, et al. Characterizing human cytomegalovirus reinfection in congenitally infected infants: an evolutionary perspective. Mol Ecol. 2017;26(7):1980–90. Epub 2016/12/19. doi: 10.1111/mec.13953. PubMed PMID: 27988973.

19. Sackman AM, Pfeifer SP, Kowalik TF, Jensen JD. On the Demographic and Selective Forces Shaping Patterns of Human Cytomegalovirus Variation within Hosts. Pathogens. 2018;7(1). Epub 2018/02/01. doi: 10.3390/pathogens7010016. PubMed PMID: 29382090; PubMed Central PMCID: PMCPMC5874742.

20. Cudini J, Roy S, Houldcroft CJ, Bryant JM, Depledge DP, Tutill H, et al. Human cytomegalovirus haplotype reconstruction reveals high diversity due to superinfection and evidence of within-host recombination. Proc Natl Acad Sci U S A. 2019;116(12):5693–8. Epub 2019/03/02. doi: 10.1073/pnas.1818130116. PubMed PMID: 30819890; PubMed Central PMCID: PMCPMC6431178.

21. Drake AL, Roxby AC, Ongecha-Owuor F, Kiarie J, John-Stewart G, Wald A, et al. Valacyclovir suppressive therapy reduces plasma and breast milk HIV-1 RNA levels during pregnancy and postpartum: a randomized trial. J Infect Dis. 2012;205(3):366–75. Epub 2011/12/08. doi: 10.1093/infdis/jir766. PubMed PMID: 22147786; PubMed Central PMCID: PMCPMC3256951.

22. Roxby AC, Atkinson C, Asbjornsdottir K, Farquhar C, Kiarie JN, Drake AL, et al. Maternal valacyclovir and infant cytomegalovirus acquisition: a randomized controlled trial among HIV-infected women. PloS one. 2014;9(2):e87855. doi: 10.1371/journal.pone.0087855. PubMed PMID: 24504006; PubMed Central PMCID: PMC3913686.

23. Slyker J, Farquhar C, Atkinson C, Asbjornsdottir K, Roxby A, Drake A, et al. Compartmentalized cytomegalovirus replication and transmission in the setting of maternal HIV-1 infection. Clinical infectious diseases : an official publication of the Infectious Diseases Society of America. 2014;58(4):564–72. doi: 10.1093/cid/cit727. PubMed PMID: 24192386; PubMed Central PMCID: PMCPMC3905754.

24. Suarez NM, Musonda KG, Escriva E, Njenga M, Agbueze A, Camiolo S, et al. Multiple-Strain Infections of Human Cytomegalovirus With High Genomic Diversity Are Common in Breast Milk From Human Immunodeficiency Virus-Infected Women in Zambia. J Infect Dis. 2019;220(5):792–801. Epub 2019/05/06. doi: 10.1093/infdis/jiz209. PubMed PMID: 31050737; PubMed Central PMCID: PMCPMC6667993.

25. Goldstein RA, Tamuri AU, Roy S, Breuer J. Haplotype assignment of virus NGS data using co-variation of variant frequencies. bioRxiv. 2018. doi: https://doi.org/10.1101/444877.

26. Renzette N, Bhattacharjee B, Jensen JD, Gibson L, Kowalik TF. Extensive genome-wide variability of human cytomegalovirus in congenitally infected infants. PLoS Pathog. 2011;7(5):e1001344. Epub 2011/06/01. doi: 10.1371/journal.ppat.1001344. PubMed PMID: 21625576; PubMed Central PMCID: PMCPMC3098220.

27. Puchhammer-Stockl E, Gorzer I, Zoufaly A, Jaksch P, Bauer CC, Klepetko W, et al. Emergence of multiple cytomegalovirus strains in blood and lung of lung transplant recipients. Transplantation. 2006;81(2):187–94. Epub 2006/01/27. doi: 10.1097/01.tp.0000194858.50812.cb. PubMed PMID: 16436961.

28. Hage E, Wilkie GS, Linnenweber-Held S, Dhingra A, Suarez NM, Schmidt JJ, et al. Characterization of Human Cytomegalovirus Genome Diversity in Immunocompromised Hosts by Whole-Genome Sequencing Directly From Clinical Specimens. J Infect Dis. 2017;215(11):1673–83. Epub 2017/04/04. doi: 10.1093/infdis/jix157. PubMed PMID: 28368496.

29. Kadambari S, Atkinson C, Luck S, Macartney M, Conibear T, Harrison I, et al. Characterising variation in five genetic loci of cytomegalovirus during treatment for congenital infection. J Med Virol. 2017;89(3):502–7. Epub 2016/08/04. doi: 10.1002/jmv.24654. PubMed PMID: 27486960.

30. Ross SA, Novak Z, Pati S, Patro RK, Blumenthal J, Danthuluri VR, et al. Mixed infection and strain diversity in congenital cytomegalovirus infection. J Infect Dis. 2011;204(7):1003–7. Epub 2011/09/02. doi: 10.1093/infdis/jir457. PubMed PMID: 21881114; PubMed Central PMCID: PMCPMC3164425.

31. Renzette N, Gibson L, Bhattacharjee B, Fisher D, Schleiss MR, Jensen JD, et al. Rapid intrahost evolution of human cytomegalovirus is shaped by demography and positive selection. PLoS Genet. 2013;9(9):e1003735. doi: 10.1371/journal.pgen.1003735. PubMed PMID: 24086142; PubMed Central PMCID: PMCPMC3784496.

32. Vera Cruz D, Nelson CS, Tran D, Barry PA, Kaur A, Koelle K, et al. Intrahost cytomegalovirus population genetics following antibody pretreatment in a monkey model of congenital transmission. PLoS Pathog. 2020;16(2):e1007968. Epub 2020/02/15. doi: 10.1371/journal.ppat.1007968. PubMed PMID: 32059027.

33. Stanton RJ, Baluchova K, Dargan DJ, Cunningham C, Sheehy O, Seirafian S, et al. Reconstruction of the complete human cytomegalovirus genome in a BAC reveals RL13 to be a potent inhibitor of replication. J Clin Invest. 2010;120(9):3191–208. Epub 2010/08/04. doi: 10.1172/JCI42955. PubMed PMID: 20679731; PubMed Central PMCID: PMCPMC2929729.

34. Hansen SG, Powers CJ, Richards R, Ventura AB, Ford JC, Siess D, et al. Evasion of CD8+ T cells is critical for superinfection by cytomegalovirus. Science. 2010;328(5974):102–6. Epub 2010/04/03. doi: 10.1126/science.1185350. PubMed PMID: 20360110; PubMed Central PMCID: PMCPMC2883175.

35. Wang L, Xu X, Zhang H, Qian J, Zhu J. Dried blood spots PCR assays to screen congenital cytomegalovirus infection: a meta-analysis. Virol J. 2015;12:60. doi: 10.1186/s12985-015-0281-9. PubMed PMID: 25889596; PubMed Central PMCID: PMCPMC4408583.

36. Mayer BT, Krantz EM, Swan D, Ferrenberg J, Simmons K, Selke S, et al. Transient Oral Human Cytomegalovirus Infections Indicate Inefficient Viral Spread from Very Few Initially Infected Cells. J Virol. 2017;91(12). doi: 10.1128/JVI.00380-17. PubMed PMID: 28381570; PubMed Central PMCID: PMCPMC5446638.

37. Joseph SB, Swanstrom R, Kashuba AD, Cohen MS. Bottlenecks in HIV-1 transmission: insights from the study of founder viruses. Nat Rev Microbiol. 2015;13(7):414–25. doi: 10.1038/nrmicro3471. PubMed PMID: 26052661; PubMed Central PMCID: PMCPMC4793885.

38. Cortese M, Calo S, D’Aurizio R, Lilja A, Pacchiani N, Merola M. Recombinant human cytomegalovirus (HCMV) RL13 binds human immunoglobulin G Fc. PloS one. 2012;7(11):e50166. Epub 2012/12/12. doi: 10.1371/journal.pone.0050166. PubMed PMID: 23226246; PubMed Central PMCID: PMCPMC3511460.

39. Van Damme E, Van Loock M. Functional annotation of human cytomegalovirus gene products: an update. Front Microbiol. 2014;5:218. Epub 2014/06/07. doi: 10.3389/fmicb.2014.00218. PubMed PMID: 24904534; PubMed Central PMCID: PMCPMC4032930.

40. Perez-Carmona N, Martinez-Vicente P, Farre D, Gabaev I, Messerle M, Engel P, et al. A Prominent Role of the Human Cytomegalovirus UL8 Glycoprotein in Restraining Proinflammatory Cytokine Production by Myeloid Cells at Late Times during Infection. J Virol. 2018;92(9). Epub 2018/02/23. doi: 10.1128/JVI.02229-17. PubMed PMID: 29467314; PubMed Central PMCID: PMCPMC5899185.

41. Bruno L, Cortese M, Monda G, Gentile M, Calo S, Schiavetti F, et al. Human cytomegalovirus pUL10 interacts with leukocytes and impairs TCR-mediated T-cell activation. Immunol Cell Biol. 2016;94(9):849–60. Epub 2016/10/19. doi: 10.1038/icb.2016.49. PubMed PMID: 27192938.

42. Gabaev I, Elbasani E, Ameres S, Steinbruck L, Stanton R, Doring M, et al. Expression of the human cytomegalovirus UL11 glycoprotein in viral infection and evaluation of its effect on virus-specific CD8 T cells. J Virol. 2014;88(24):14326–39. Epub 2014/10/03. doi: 10.1128/JVI.01691-14. PubMed PMID: 25275132; PubMed Central PMCID: PMCPMC4249143.

43. Pereira L, Tabata T, Petitt M, Fang-Hoover J. Congenital cytomegalovirus infection undermines early development and functions of the human placenta. Placenta. 2017;59 Suppl 1:S8–S16. Epub 2017/05/10. doi: 10.1016/j.placenta.2017.04.020. PubMed PMID: 28477968.

44. Frank T, Niemann I, Reichel A, Stamminger T. Emerging roles of cytomegalovirus-encoded G protein-coupled receptors during lytic and latent infection. Med Microbiol Immunol. 36. 2019;208(3-4):447–56. Epub 2019/03/23. doi: 10.1007/s00430-019-00595-9. PubMed PMID: 30900091.

45. Heatley SL, Pietra G, Lin J, Widjaja JM, Harpur CM, Lester S, et al. Polymorphism in human cytomegalovirus UL40 impacts on recognition of human leukocyte antigen-E (HLA-E) by natural killer cells. J Biol Chem. 2013;288(12):8679–90. Epub 2013/01/22. doi: 10.1074/jbc.M112.409672. PubMed PMID: 23335510; PubMed Central PMCID: PMCPMC3605686.

46. Lee MK, Kim YJ, Kim YE, Han TH, Milbradt J, Marschall M, et al. Transmembrane Protein pUL50 of Human Cytomegalovirus Inhibits ISGylation by Downregulating UBE1L. J Virol. 2018;92(15). Epub 2018/05/11. doi: 10.1128/JVI.00462-18. PubMed PMID: 29743376; PubMed Central PMCID: PMCPMC6052311.

47. DeRussy BM, Boland MT, Tandon R. Human Cytomegalovirus pUL93 Links Nucleocapsid Maturation and Nuclear Egress. J Virol. 2016;90(16):7109–17. Epub 2016/05/27. doi: 10.1128/JVI.00728-16. PubMed PMID: 27226374; PubMed Central PMCID: PMCPMC4984640.

48. Wu Y, Prager A, Boos S, Resch M, Brizic I, Mach M, et al. Human cytomegalovirus glycoprotein complex gH/gL/gO uses PDGFR-alpha as a key for entry. PLoS Pathog. 2017;13(4):e1006281. Epub 2017/04/14. doi: 10.1371/journal.ppat.1006281. PubMed PMID: 28403202; PubMed Central PMCID: PMCPMC5389851.

49. Zischke J, Mamareli P, Pokoyski C, Gabaev I, Buyny S, Jacobs R, et al. The human cytomegalovirus glycoprotein pUL11 acts via CD45 to induce T cell IL-10 secretion. PLoS Pathog. 2017;13(6):e1006454. Epub 2017/06/20. doi: 10.1371/journal.ppat.1006454. PubMed PMID: 28628650; PubMed Central PMCID: PMCPMC5491327.

50. Gatherer D, Seirafian S, Cunningham C, Holton M, Dargan DJ, Baluchova K, et al. High-resolution human cytomegalovirus transcriptome. Proc Natl Acad Sci U S A. 2011;108(49):19755–60. Epub 2011/11/24. doi: 10.1073/pnas.1115861108. PubMed PMID: 22109557; PubMed Central PMCID: PMCPMC3241806.

51. Lurain NS, Fox AM, Lichy HM, Bhorade SM, Ware CF, Huang DD, et al. Analysis of the human cytomegalovirus genomic region from UL146 through UL147A reveals sequence hypervariability, genotypic stability, and overlapping transcripts. Virol J. 2006;3:4. Epub 2006/01/18. doi: 10.1186/1743-422X-3-4. PubMed PMID: 16409621; PubMed Central PMCID: PMCPMC1360065.

52. Arav-Boger R, Foster CB, Zong JC, Pass RF. Human cytomegalovirus-encoded alpha - chemokines exhibit high sequence variability in congenitally infected newborns. J Infect Dis. 2006;193(6):788–91. Epub 2006/02/16. doi: 10.1086/500508. PubMed PMID: 16479512.

53. Arcangeletti MC, Vasile Simone R, Rodighiero I, De Conto F, Medici MC, Martorana D, et al. Combined genetic variants of human cytomegalovirus envelope glycoproteins as congenital infection markers. Virol J. 2015;12:202. Epub 2015/11/28. doi: 10.1186/s12985-015-0428-8. PubMed PMID: 26611326; PubMed Central PMCID: PMCPMC4662005.

54. Houldcroft CJ, Bryant JM, Depledge DP, Margetts BK, Simmonds J, Nicolaou S, et al. Detection of Low Frequency Multi-Drug Resistance and Novel Putative Maribavir Resistance in Immunocompromised Pediatric Patients with Cytomegalovirus. Front Microbiol. 2016;7:1317. Epub 2016/09/27. doi: 10.3389/fmicb.2016.01317. PubMed PMID: 27667983; PubMed Central PMCID: PMCPMC5016526.

55. Katoh K, Standley DM. MAFFT multiple sequence alignment software version 7: improvements in performance and usability. Mol Biol Evol. 2013;30(4):772–80. Epub 2013/01/19. doi: 10.1093/molbev/mst010. PubMed PMID: 23329690; PubMed Central PMCID: PMCPMC3603318.

56. Paradis E, Schliep K. ape 5.0: an environment for modern phylogenetics and evolutionary analyses in R. Bioinformatics. 2019;35(3):526–8. Epub 2018/07/18. doi: 10.1093/bioinformatics/bty633. PubMed PMID: 30016406.

57. Team RC. A language and environment for statistical computing. R Foundation for Statistical Computing. Vienna, Austria 2012.

58. Akaike H. Information theory and an extension of the maximum likelihood principle. 2nd International Symposium on Information Theory (BN Petrov and F Csäki, eds); Akademiai Ki à do, Budapest 1973.

59. Stamatakis A. RAxML version 8: a tool for phylogenetic analysis and post-analysis of large phylogenies. Bioinformatics. 2014;30(9):1312–3. Epub 2014/01/24. doi: 10.1093/bioinformatics/btu033. PubMed PMID: 24451623; PubMed Central PMCID: PMCPMC3998144.

